# Genetically nominated cardiovascular-kidney-metabolic features are not consistently more responsive to weight-loss and cardiometabolic interventions

**DOI:** 10.64898/2026.07.27.26358659

**Authors:** Cheng-Yu Hsueh, Ko-Ting Chen, Bertrand Chin-Ming Tan

## Abstract

**Background:** Substantial cardiovascular and kidney risk persists after successful weight loss. Whether features that genetics implicates in cardiovascular-kidney-metabolic (CKM) disease show larger short-term responses to weight-loss and cardiometabolic interventions than other features is unknown.

**Methods:** Across four molecular layers (proteome, transcriptome, methylome, metabolome), we applied layer-specific cis-Mendelian randomization with colocalization or summary-data shared-signal filtering against eight CKM genome-wide association studies. Within 13 omics-by-intervention analyses (diet, bariatric surgery, empagliflozin, behavioral weight loss), we compared response magnitude between nominated and adequately-instrumented non-nominated features using rank-based Cliff’s δ; methylation was baseline-variance-matched and directional analyses exploratory.

**Results:** Here we show genetic nomination is not consistently associated with larger observed response magnitude: estimates are small (|δ| < 0.08 in all 13; median |δ| = 0.03), near zero (descriptive pooled δ = +0.001, 95% CI −0.013 to +0.015; I² = 0%), none significant by label-permutation, though smaller proteomic and transcriptomic analyses remain compatible with modest differences. Nominated proteins and metabolites frequently changed (48–71% were responsive), at rates similar to comparison features; the null is not one of universal non-response. Nominated CpG sites have lower baseline inter-individual variance (Mann–Whitney *P* = 1.7 × 10⁻⁶ to 4 × 10⁻³ at the primary nomination tiers), and apparent methylation persistence attenuates after variance-matching.

**Conclusions:** Under the definitions and datasets studied, genetic target support does not consistently predict pharmacodynamic responsiveness and should not be read as a treatment-response or reversibility biomarker; baseline dynamic range should be assessed and controlled when comparing molecular change across selected features. Nominated features are not shown unchanged or to explain residual risk; same-subject longitudinal studies are needed to test prognostic or therapeutic value.

**Plain Language Summary:** People who lose weight often see their heart, kidney and metabolic health improve, yet some risk remains. We asked whether the molecular features that genetics links to these diseases are also the ones that improve most with treatment. We gathered four kinds of molecular measurement from studies of dieting, weight-loss surgery and medicines, and compared them with other measured molecules. They still changed with treatment — but no more than the rest, with no consistent link to their genetic importance. An apparent lasting “memory” of obesity largely reflected how the molecules were measured rather than a persistent biological change. On this evidence, genetically implicated features do not consistently respond more, and better-designed studies are needed.

## Introduction

Weight loss is the cornerstone of treatment for cardiovascular-kidney-metabolic disease, and intentional loss improves glycemia, blood pressure, liver fat and albuminuria [1]. Yet residual cardiovascular and kidney events remain common after successful weight loss, including in the contemporary trials of incretin and bariatric therapies [2,3]. Which of the many molecular changes that accompany weight loss mark disease-driving biology, rather than tracking the loss of fat mass itself, is unresolved. Distinguishing the two is a prerequisite for reading a molecular response as evidence that a disease-relevant target has been engaged.

Weight-loss and cardiometabolic interventions change thousands of molecular features across the proteome, transcriptome, methylome and metabolome [4]. Many such changes are downstream or state-dependent correlates of adiposity rather than features on the causal path to disease. Many weight-loss-associated protein changes, for example, are attributable to the change in body-mass index [4]. Human genetic evidence offers an independent way to prioritize disease-relevant features. Genetically supported drug targets are roughly 2.6-fold more likely to succeed in clinical development [5]. Cis-acting Mendelian randomization combined with colocalization against disease genome-wide association studies (GWAS) is an established framework for nominating candidate disease-relevant molecular features. It also guards against confounding by linkage disequilibrium, which invalidates roughly a third of naïve Mendelian-randomization signals [6].

These two kinds of evidence index different things. Genetic nomination reflects a feature’s lifelong, germline-set association with disease — an exposure randomized at conception and therefore largely free of reverse causation and confounding [7]. Intervention response reflects a feature’s acquired, state-dependent plasticity. The two need not coincide. A feature can be a faithful pharmacodynamic marker of an intervention’s activity without lying on the causal path to disease. Conversely, a genetically nominated causal feature need not be the one that moves most in the short term. Whether genetically nominated CKM features respond to weight-loss and cardiometabolic interventions more than other measured features has not been tested with genetic nomination as the entry filter.

Two single-layer findings frame the question. In adipose tissue, obesity-associated transcriptional changes persist after weight loss in human and mouse, and epigenomic changes persist in mouse adipocytes — an “epigenetic memory of obesity” [8]. Separately, reverse-direction Mendelian randomization in the transcriptome indicates that most disease-associated differentially expressed genes reflect reactive rather than causal changes [9]. Neither study used genetic nomination as a uniform entry filter, neither spanned more than one molecular layer, and neither tested the same gene across layers. Our human bulk DNA-methylation test of persistence is correspondingly narrower than the mouse-adipocyte epigenome of Hinte et al. [8].

We therefore asked one question across the molecular cascade: do features genetically nominated for CKM association show larger molecular responses to weight-loss and cardiometabolic interventions than non-nominated features? We classified every adequately-instrumented feature in four layers as genetically nominated for CKM association or non-nominated, using layer-specific cis-QTL Mendelian randomization with shared-signal or pleiotropy filtering against eight CKM-disease GWAS. We then compared the two groups on two axes: response magnitude (the primary endpoint) and restorative direction (a secondary, exploratory axis). The analysis was designed as an unbiased, layer-wide screen rather than a re-test of prior hit lists. Leave-eGFR-out and per-outcome-balanced analyses were pre-specified to prevent any single well-powered outcome from dominating the nominated set. Across all four layers, genetic nomination was not consistently associated with a greater molecular response. An apparent methylation “memory” signal was sensitive to differences in baseline variance rather than indicating preferential persistence of nominated marks.

## Results

### A uniform genetics-anchored framework for comparing nominated and non-nominated features

We built one conceptual framework and implemented it with layer-specific nomination pipelines across four layers. For each measured feature — a protein, a transcript, a CpG site or a metabolite — we tested whether its cis-genetic instruments supported an effect on each of eight CKM outcomes by layer-specific cis-QTL Mendelian randomization with a shared-signal or pleiotropy filter (Methods). The eight outcomes were type 2 diabetes, coronary artery disease, atrial fibrillation, heart failure, stroke, chronic kidney disease, estimated glomerular filtration rate and non-alcoholic fatty liver disease. We labeled the feature genetically nominated for CKM association or non-nominated. We then measured the same feature’s molecular response in paired human intervention data. We compared response magnitude between nominated and non-nominated features, and whether the response moved opposite to the feature’s disease-associated direction (restorative). Nominations drew on established cis-QTL atlases: pQTL (UKB-PPP, deCODE), eQTL (GTEx via the eQTL Catalogue), mQTL (GoDMC) and metaboQTL (Chen-CLSA). Response drew on paired diet, bariatric, incretin, SGLT2-inhibitor and exercise data (Table 1). Features were harmonized by direction only, never by raw effect magnitude, because the layers are measured on non-comparable scales. The comparison was restricted to adequately-instrumented features, to reduce differential opportunity for nomination arising solely from the absence of an instrument. Residual differences between the groups in QTL coverage, tissue relevance and disease-GWAS power remain.

**Table 1.**
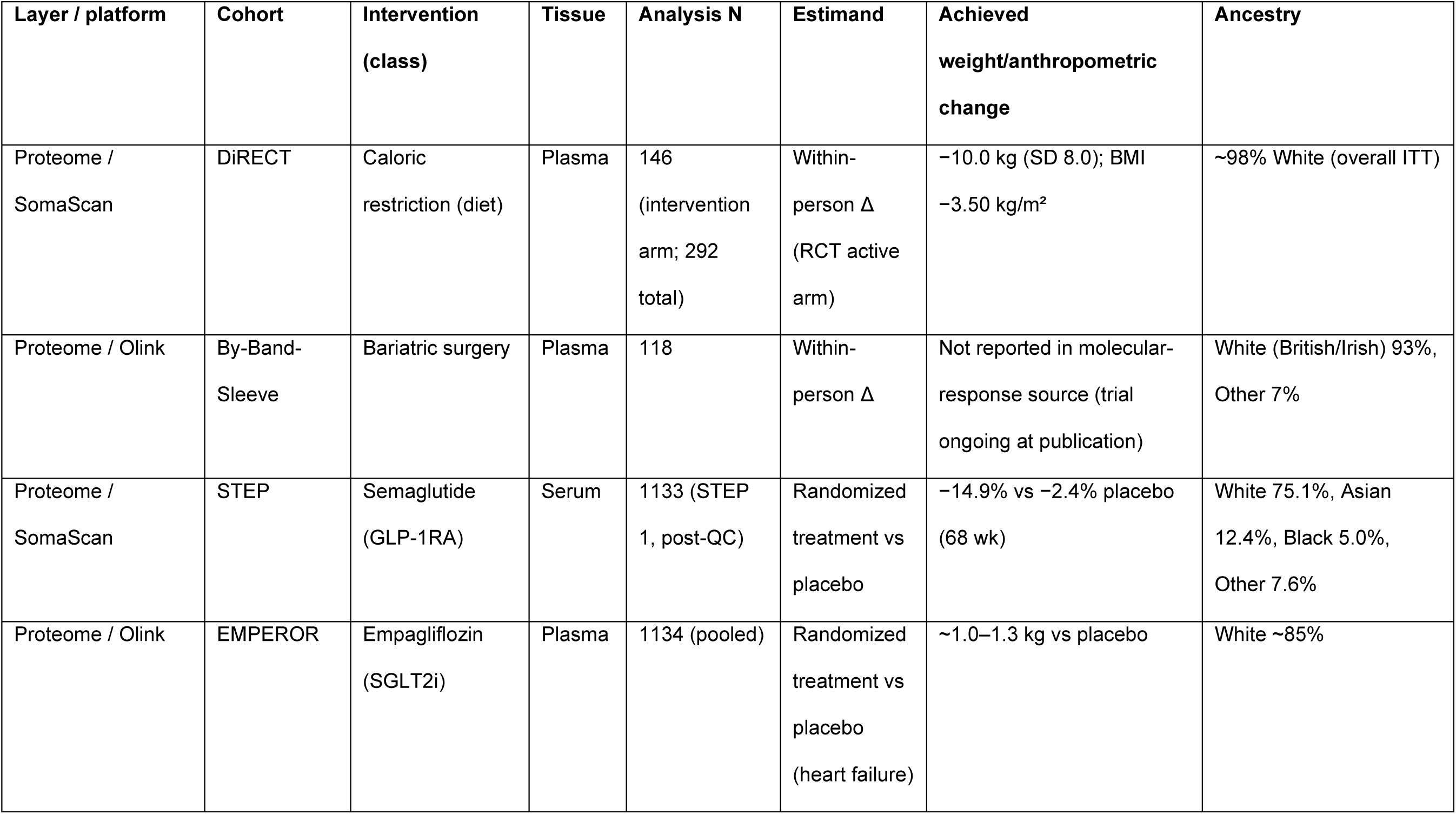
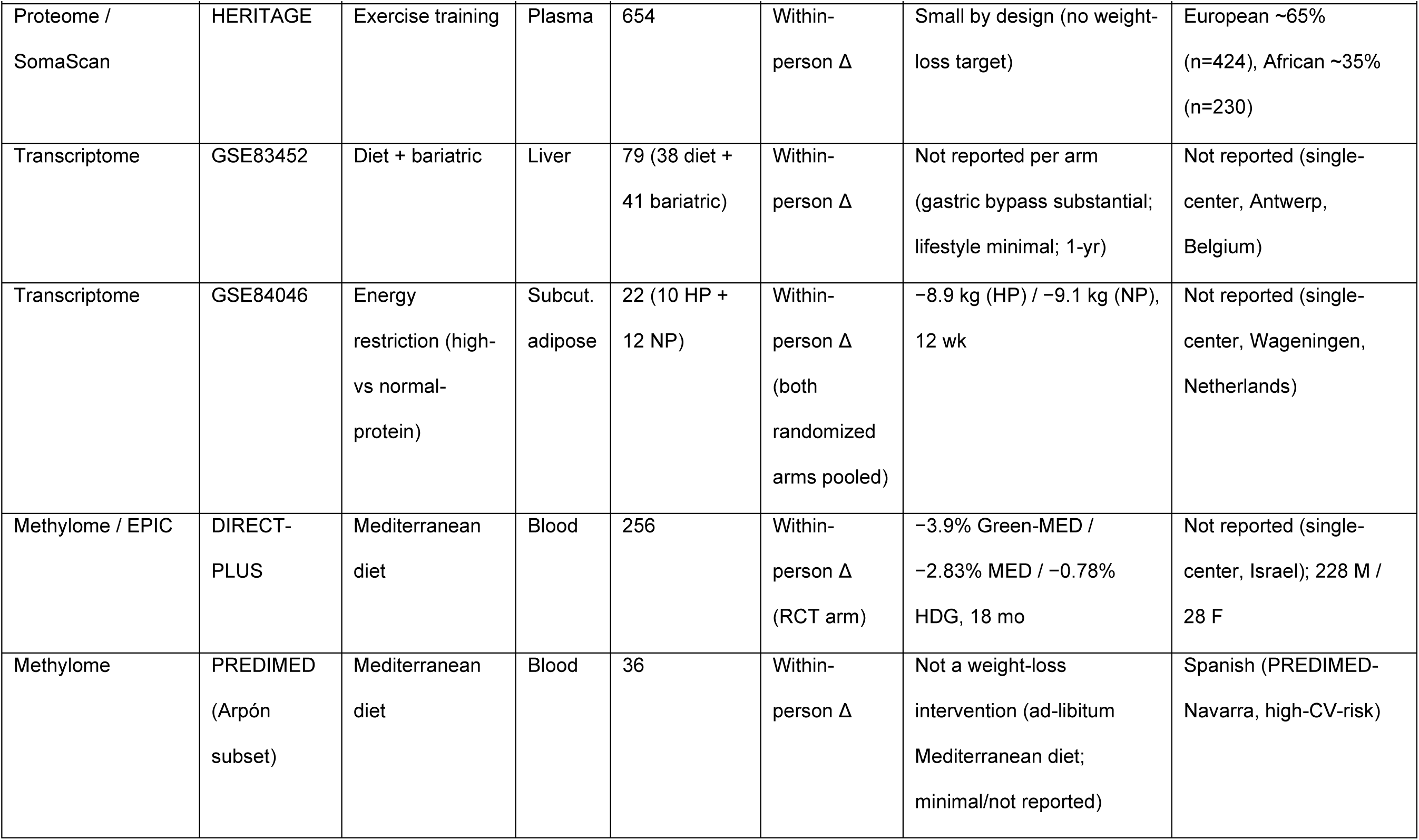
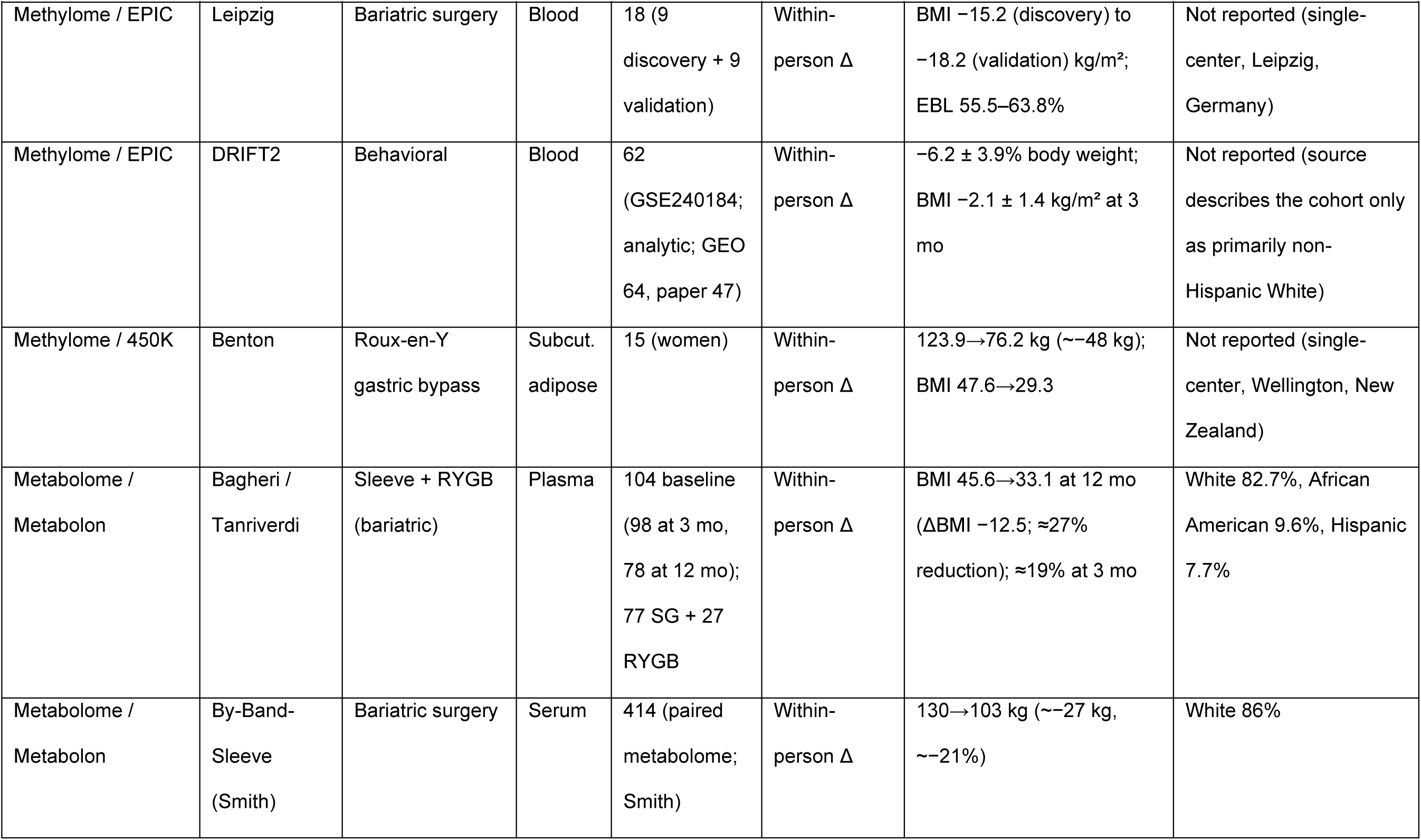
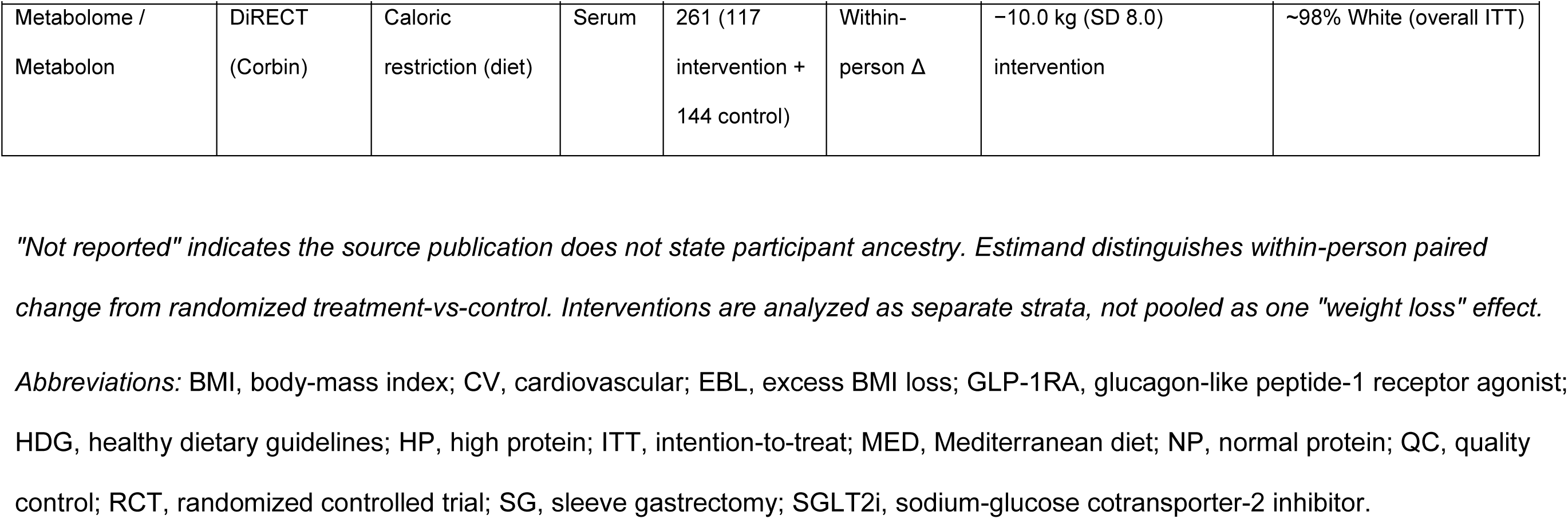
Study characteristics — cohorts, interventions, and intervention data.

| Layer / platform | Cohort | Intervention<br>(class) | Tissue | Analysis N | Estimand | Achieved<br>weight/anthropometric<br>change | Ancestry |
| --- | --- | --- | --- | --- | --- | --- | --- |
| Proteome /<br>SomaScan | DiRECT | Caloric<br>restriction (diet) | Plasma | 146<br>(intervention<br>arm; 292<br>total) | Within-person $\Delta$<br>(RCT active<br>arm) | −10.0 kg (SD 8.0); BMI<br>−3.50 kg/m<sup>2</sup> | ~98% White (overall ITT) |
| Proteome / Olink | By-Band-Sleeve | Bariatric surgery | Plasma | 118 | Within-person $\Delta$ | Not reported in molecular-response source (trial<br>ongoing at publication) | White (British/Irish) 93%,<br>Other 7% |
| Proteome /<br>SomaScan | STEP | Semaglutide<br>(GLP-1RA) | Serum | 1133 (STEP<br>1, post-QC) | Randomized<br>treatment vs<br>placebo | −14.9% vs −2.4% placebo<br>(68 wk) | White 75.1%, Asian<br>12.4%, Black 5.0%,<br>Other 7.6% |
| Proteome / Olink | EMPEROR | Empagliflozin<br>(SGLT2i) | Plasma | 1134 (pooled) | Randomized<br>treatment vs<br>placebo<br>(heart failure) | ~1.0–1.3 kg vs placebo | White ~85% |

| Layer / platform | Cohort | Intervention (class) | Tissue | Analysis N | Estimand | Achieved weight/anthropometric change | Ancestry |
| --- | --- | --- | --- | --- | --- | --- | --- |
| Proteome / SomaScan | HERITAGE | Exercise training | Plasma | 654 | Within-person $\Delta$ | Small by design (no weight-loss target) | European ~65% (n=424), African ~35% (n=230) |
| Transcriptome | GSE83452 | Diet + bariatric | Liver | 79 (38 diet + 41 bariatric) | Within-person $\Delta$ | Not reported per arm (gastric bypass substantial; lifestyle minimal; 1-yr) | Not reported (single-center, Antwerp, Belgium) |
| Transcriptome | GSE84046 | Energy restriction (high-vs normal-protein) | Subcut. adipose | 22 (10 HP + 12 NP) | Within-person $\Delta$ (both randomized arms pooled) | -8.9 kg (HP) / -9.1 kg (NP), 12 wk | Not reported (single-center, Wageningen, Netherlands) |
| Methylome / EPIC | DIRECT-PLUS | Mediterranean diet | Blood | 256 | Within-person $\Delta$ (RCT arm) | -3.9% Green-MED / -2.83% MED / -0.78% HDG, 18 mo | Not reported (single-center, Israel); 228 M / 28 F |
| Methylome | PREDIMED (Arpón subset) | Mediterranean diet | Blood | 36 | Within-person $\Delta$ | Not a weight-loss intervention (ad-libitum Mediterranean diet; minimal/not reported) | Spanish (PREDIMED-Navarra, high-CV-risk) |
| Methylome / EPIC | Leipzig | Bariatric surgery | Blood | 18 (9<br>discovery + 9<br>validation) | Within-person $\Delta$ | BMI -15.2 (discovery) to<br>-18.2 (validation) kg/m<sup>2</sup>;<br>EBL 55.5–63.8% | Not reported (single-center, Leipzig,<br>Germany) |
| Methylome / EPIC | DRIFT2 | Behavioral | Blood | 62<br>(GSE240184;<br>analytic; GEO<br>64, paper 47) | Within-person $\Delta$ | -6.2 ± 3.9% body weight;<br>BMI -2.1 ± 1.4 kg/m<sup>2</sup> at 3<br>mo | Not reported (source<br>describes the cohort only<br>as primarily non-Hispanic White) |
| Methylome / 450K | Benton | Roux-en-Y<br>gastric bypass | Subcut.<br>adipose | 15 (women) | Within-person $\Delta$ | 123.9→76.2 kg (~-48 kg);<br>BMI 47.6→29.3 | Not reported (single-center, Wellington, New<br>Zealand) |
| Metabolome /<br>Metabolon | Bagheri /<br>Tanriverdi | Sleeve + RYGB<br>(bariatric) | Plasma | 104 baseline<br>(98 at 3 mo,<br>78 at 12 mo);<br>77 SG + 27<br>RYGB | Within-person $\Delta$ | BMI 45.6→33.1 at 12 mo<br>(ΔBMI -12.5; ≈27%<br>reduction); ≈19% at 3 mo | White 82.7%, African<br>American 9.6%, Hispanic<br>7.7% |
| Metabolome /<br>Metabolon | By-Band-Sleeve<br>(Smith) | Bariatric surgery | Serum | 414 (paired<br>metabolome;<br>Smith) | Within-person $\Delta$ | 130→103 kg (~-27 kg,<br>~-21%) | White 86% |
| Metabolome /<br>Metabolon | DiRECT<br>(Corbin) | Caloric<br>restriction (diet) | Serum | 261 (117<br>intervention +<br>144 control) | Within-person $\Delta$ | -10.0 kg (SD 8.0)<br>intervention | ~98% White (overall ITT) |

*"Not reported" indicates the source publication does not state participant ancestry. Estimand distinguishes within-person paired*
*change from randomized treatment-vs-control. Interventions are analyzed as separate strata, not pooled as one "weight loss" effect.*
*Abbreviations:* BMI, body-mass index; CV, cardiovascular; EBL, excess BMI loss; GLP-1RA, glucagon-like peptide-1 receptor agonist;
HDG, healthy dietary guidelines; HP, high protein; ITT, intention-to-treat; MED, Mediterranean diet; NP, normal protein; QC, quality
control; RCT, randomized controlled trial; SG, sleeve gastrectomy; SGLT2i, sodium-glucose cotransporter-2 inhibitor.

One property of the nominated sets required correction throughout. The number of features nominated tracked the statistical power of the outcome GWAS. Estimated glomerular filtration rate, the only quantitative trait and the best-powered GWAS, dominated every layer’s nominated set. Nominated-set size varied markedly across the eight outcomes, and eGFR — the only quantitative trait among them — yielded by far the largest nominated set (Supplementary Table S3, Supplementary Fig. S3), consistent with differential power. We therefore pre-specified leave-eGFR-out and per-outcome-balanced analyses as the primary contrasts for the proteome, transcriptome and metabolome, and report the eGFR-inclusive sets as secondary. For the methylome, baseline inter-individual variance rather than outcome-GWAS power is the dominant confound. The headline contrast is instead the variance-matched eGFR-inclusive analysis, with leave-eGFR-out as a sensitivity check (Methods). Established disease genes were recovered (PCSK9, IL6R and APOB for coronary disease; GCKR for type 2 diabetes; Fig. 1c), indicating sensitivity where strong causal biology exists. This calibrates sensitivity, not specificity.

**Figure 1.**
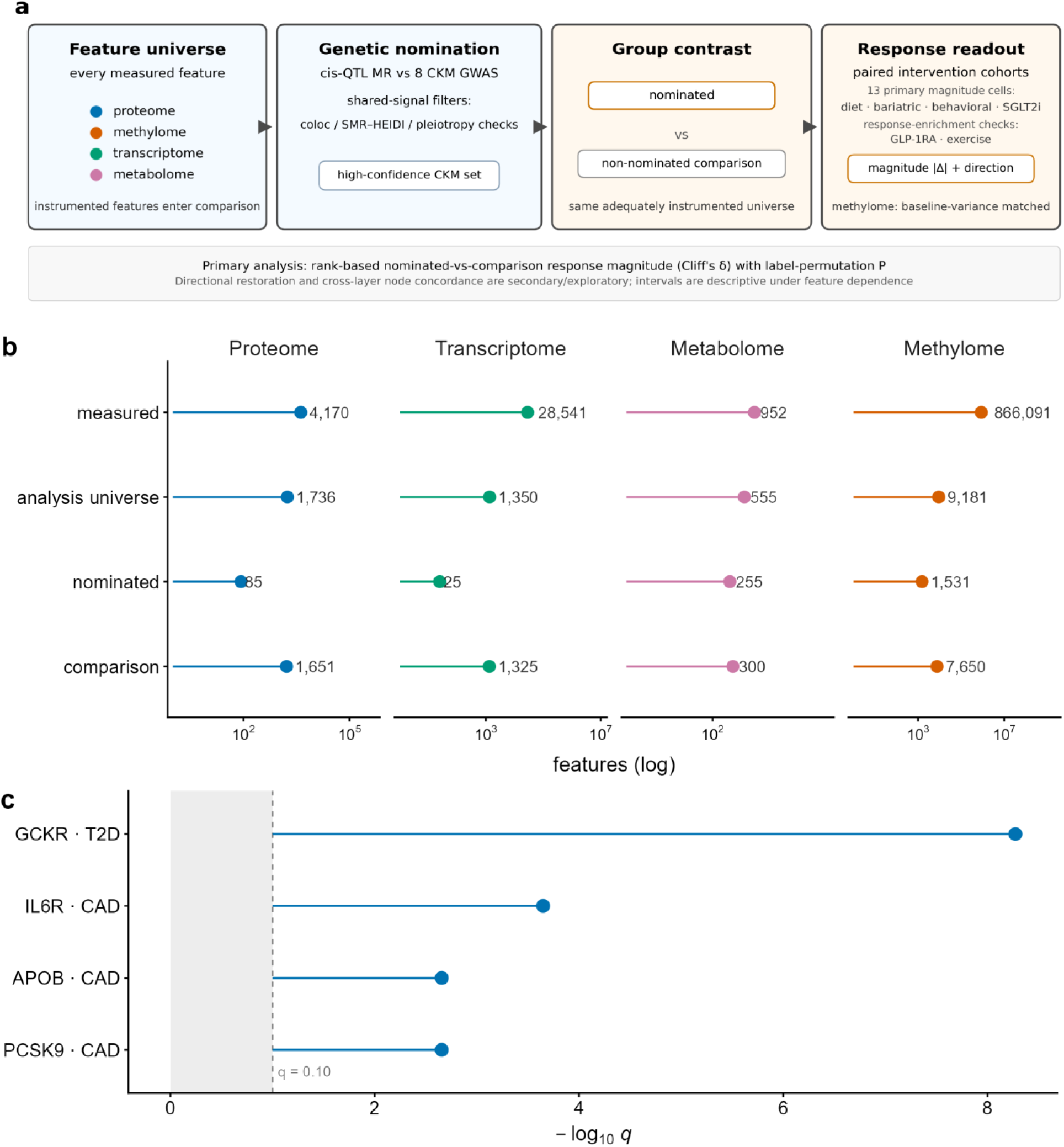
Study design, feature flow, and calibration. **(a)** Schematic of the genetics-anchored design: every measured feature in four molecular layers is classified by layer-specific cis-QTL Mendelian randomization with shared-signal or pleiotropy filtering against eight CKM GWAS (*q* < 0.10, leave-eGFR-out) as genetically nominated or non-nominated, then compared for molecular response magnitude and restorative direction across weight-loss and cardiometabolic intervention classes. The response box separates the four classes that contribute the 13 primary magnitude cells (diet, bariatric surgery, behavioral weight loss and SGLT2 inhibition) from the two interventions examined only as response-enrichment checks (glucagon-like peptide-1 receptor agonism and exercise training). **(b)** Feature flow per layer (log scale; each layer’s panel is scaled independently, so bar lengths are comparable only within a panel — read the printed counts across layers): measured (reversal-panel size) → analysis universe, the set entering each contrast (nominated + comparison) → nominated and non-nominated comparison features; the nominated/comparison counts are the sets entering each representative contrast (proteome 85/1,651; transcriptome 25/1,325; metabolome 255/300; methylome 1,531/7,650), reconciling with Figs 2–3 and Table 2 (full per-layer feature flow, Supplementary Table S1). Two unit conventions apply below the measured row: the proteome counts are SomaScan aptamers, the unit the contrast was run on (1,736 aptamers over 1,607 distinct proteins), and the methylome comparison count is the baseline-variance-matched subsample — five per nominated CpG (Methods) — so that layer’s analysis universe is much smaller than its instrumented, measured set, whereas the other three layers’ analysis universe is that set. Nomination criteria are layer-specific (colocalization for proteome and transcriptome, SMR/HEIDI for methylome, MR with pleiotropy filtering for metabolome). **(c)** Calibration: canonical causal proteins (PCSK9, APOB and IL6R for coronary disease; GCKR for type 2 diabetes) are recovered by the pipeline (−log10 *q*; all nominated, *q* < 0.10). Source data: source_data/{flow,calibration}.tsv. Figure 1a *is a study-design schematic; all data panels are rendered from frozen source data. Per-analysis n nominated, n comparison and the small-effect (descriptive) verdict are tabulated in Table 2 (per-analysis estimand is in Table 1). Supplementary figures S1–S4 are listed at the end*.

**Table 2.**
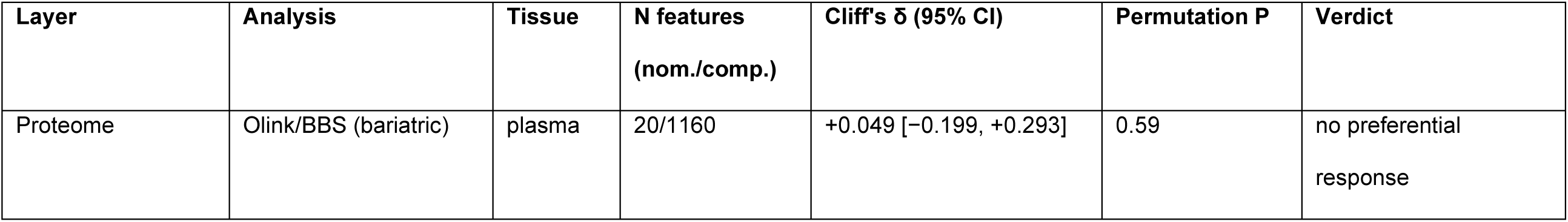
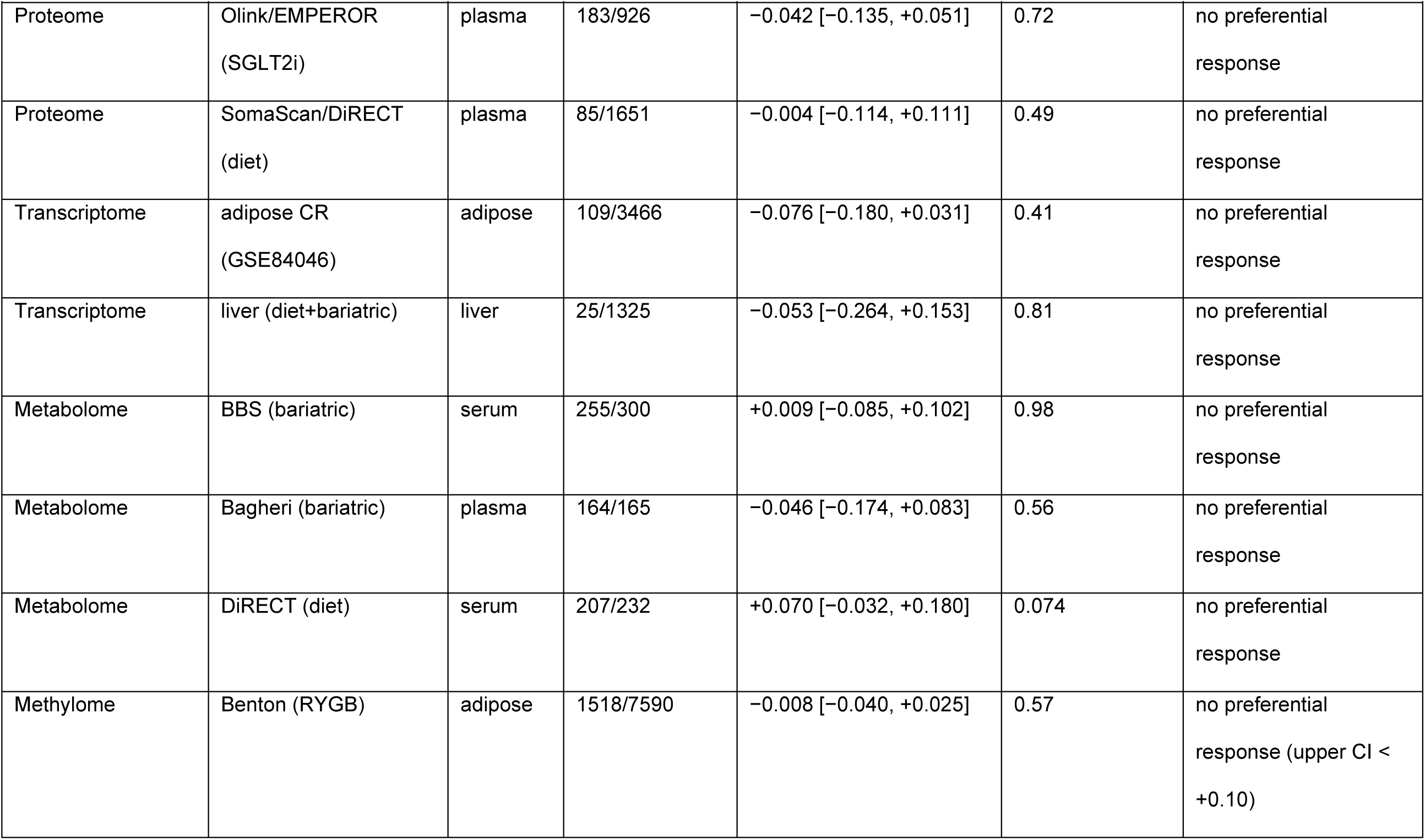
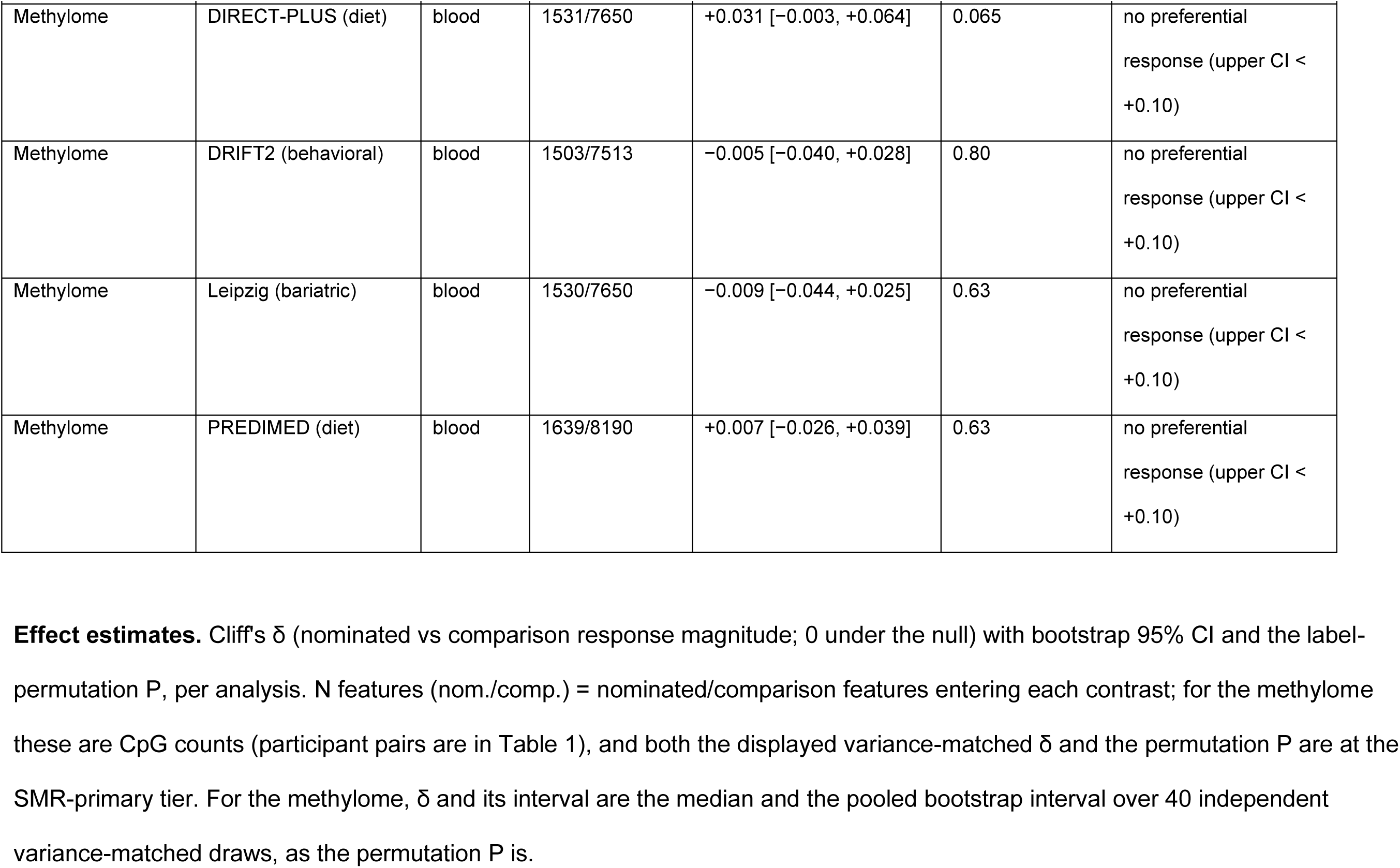
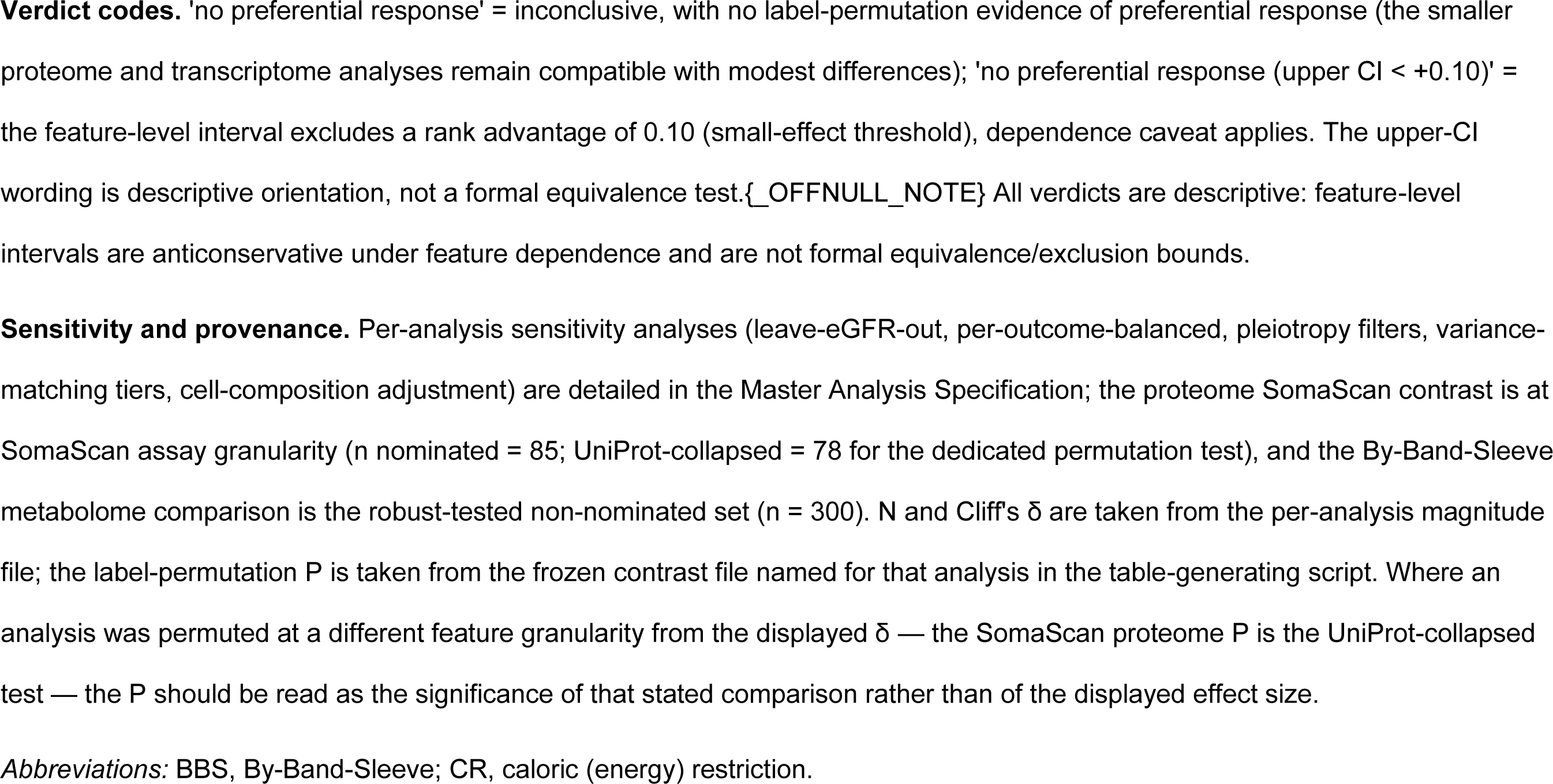
The 13 nominated-vs-comparison response-magnitude analyses.

### Genetic nomination is not consistently associated with greater molecular-response magnitude

Across 13 layer-by-intervention-by-tissue analyses, genetic nomination was not consistently associated with how much a feature responded to weight-loss and cardiometabolic interventions (Fig. 2). All 13 analyses had a Cliff’s δ whose feature-level 95% confidence interval crossed zero. Cliff’s δ is the rank advantage of nominated over comparison features, P(X_N > X_C) − P(X_N < X_C), where X_N and X_C are the response magnitudes of a nominated and a comparison feature; ties contribute to neither term, and δ is centered at zero under the null.

**Figure 2.**
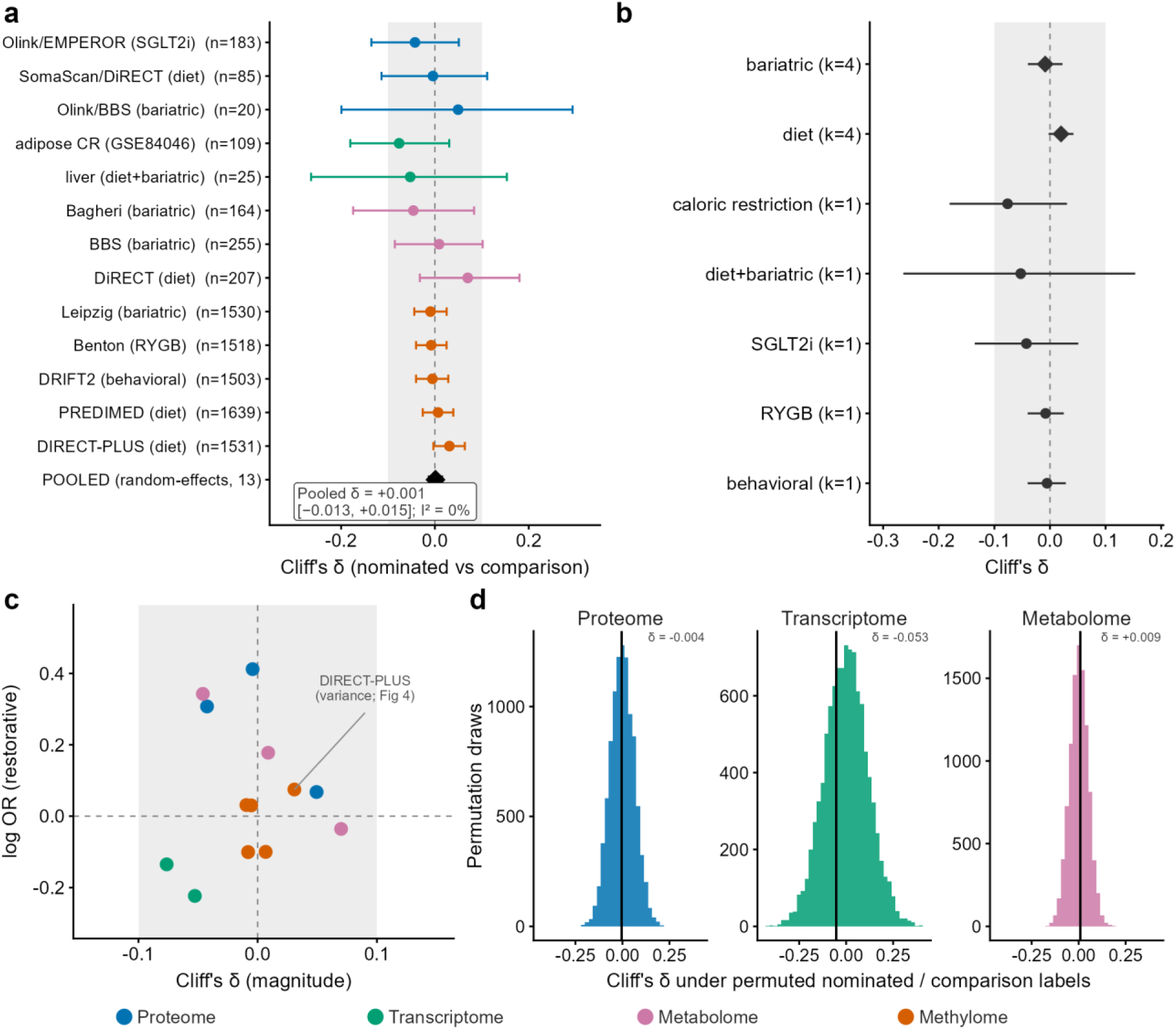
Genetic nomination is not consistently associated with greater molecular-response magnitude. **(a)** Per-analysis magnitude: Cliff’s δ (nominated versus comparison response magnitude; δ = 0 under the null) with bootstrap 95% confidence intervals for all 13 layer-by-intervention-by-tissue analyses, colored by layer, against a small-effect reference band (±0.10, gray; interpreted descriptively, not as a formal equivalence test — see Methods); the random-effects pooled estimate (diamond) is +0.001 (95% CI −0.013 to +0.015; I² = 0%) and no analysis is significant by label-permutation. For the methylome, δ and its interval are the median and the pooled bootstrap interval over 40 independent variance-matched draws, as the permutation *P* is; the cell nearest to off-null (variance-matched DIRECT-PLUS, δ = +0.031, 95% CI −0.003 to +0.064) also has a non-significant label-permutation test (*P* = 0.065 at the SMR-primary tier; Fig. 4). **(b)** The null holds within each intervention class: random-effects pooled δ per class; classes represented by a single analysis (k = 1) are shown as points rather than pooled estimates, and only diet and bariatric surgery (k = 4 each) are true pools. **(c)** Magnitude × direction: each analysis positioned by its magnitude effect (Cliff’s δ) and directional restorative effect (log odds ratio), all clustered near the null origin (dashed lines); no analysis lies off the null once the variance-matched draw is propagated into the methylome intervals (Fig. 4). **(d)** Label-permutation null: for a representative cell of the proteome, transcriptome and metabolome, the observed Cliff’s δ (vertical line) falls within the ≥10,000-permutation null (per-analysis permutation P values are given in Table 2); the methylome’s primary contrast is variance-matched and is shown in Fig. 4. Per-analysis n nominated, n comparison and the per-analysis small-effect (descriptive) verdict are in Table 2. Source data: source_data/{magnitude_effects.csv, meta_subgroup.tsv, dualnull.tsv, permutation_null.tsv}.

The primary analysis in each layer was consistent with this pattern. In the proteome, colocalization-grade nominated proteins responded no more than comparison features after bariatric surgery, in the By-Band-Sleeve Olink cohort (n = 20 nominated proteins from 19 independent loci; permutation *P* = 0.59). The result was similarly null on an independent platform, atlas and intervention — the DiRECT caloric-restriction arm, measured by SomaScan and nominated from deCODE (leave-eGFR-out: Cliff’s δ = −0.004, 95% CI −0.114 to +0.111, n = 85; a dedicated permutation test on the UniProt-aggregated set, n = 78, gave *P* = 0.49). In the transcriptome, the result held in liver (colocalization-gated n = 25; permutation *P* = 0.81) and in adipose tissue with tissue-matched adipose eQTL instruments (leave-eGFR-out n = 109; permutation *P* = 0.41). In the metabolome, nominated metabolites responded no more than comparison features in two bariatric cohorts (Bagheri leave-eGFR-out permutation *P* = 0.56; By-Band-Sleeve *P* = 0.98). In the DiRECT diet arm the leave-eGFR-out contrast was a non-significant trend in the respond-more direction (n = 207, *P* = 0.074, Cliff’s δ = +0.070). This did not persist in the primary pleiotropy-filtered tier (n = 49, *P* = 0.20). The result held under weighted-median and MR-Egger pleiotropy filtering in all three cohorts (Bagheri n = 35, *P* = 0.38; By-Band-Sleeve n = 57, *P* = 0.25; DiRECT n = 49, *P* = 0.20). The methylome is treated separately below.

The per-analysis effect estimates were small and centered near zero (|Cliff’s δ| < 0.08 in all 13 analyses, median |δ| = 0.03). No analysis was significant by label-permutation (Table 2). We report bootstrap confidence intervals but interpret them descriptively. Because molecular features within a layer are correlated and most intervention cohorts are small (median 118 participants, range 15 to 1,134; Table 1), feature-level intervals do not capture participant-level uncertainty and are anticonservative. We therefore do not read them as precise exclusion bounds. The smaller proteome and transcriptome analyses remained compatible with modest differences (Methods, Fig. 2).

The comparison did not reflect universal non-response. Nominated proteins and metabolites frequently changed after intervention: 48% to 71% responded significantly (response *P* < 0.05), similar to comparison features (49% to 67%). Genetic nomination therefore identified features that responded no more than the rest, not features the interventions failed to move (Fig. 3). The transcriptome was the exception, with nominated features responding somewhat less often (20% versus 32%). A descriptive random-effects synthesis of the 13 estimates was centered near zero (pooled Cliff’s δ = +0.001), with estimated heterogeneity of zero in this small set. Given the feature dependence above, we do not interpret its confidence interval as a precise bound (Fig. 2; the layer-stratified synthesis is Supplementary Fig. S1). A continuous analysis agreed. Among instrumented proteins, the strength of genetic evidence (−log10 of the minimum *q*) was not associated with response magnitude (Spearman ρ = −0.03, *P* = 0.32).

**Figure 3.**
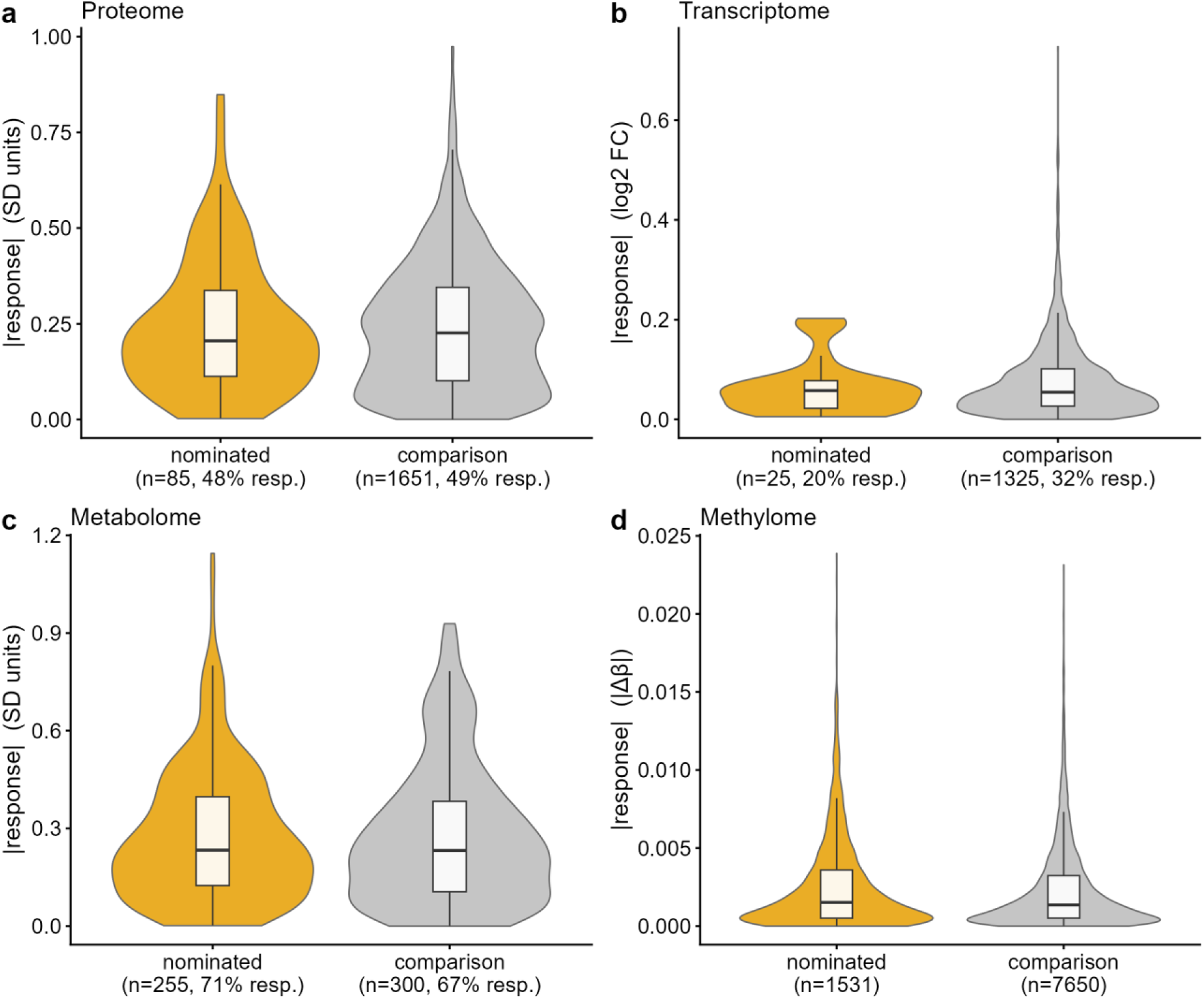
Nominated features respond similarly to comparison features. Per-feature response magnitude (|Δ|) for nominated versus comparison features, one representative dataset per layer (**a** proteome, SomaScan/DiRECT; **b** transcriptome, liver; **c** metabolome, By-Band-Sleeve; **d** methylome, DIRECT-PLUS); violins show the distribution, boxplots the median and IQR. Distributions overlap in every layer and a similar fraction of each group responds significantly (nominated versus comparison: proteome 48% versus 49%, metabolome 71% versus 67%, transcriptome 20% versus 32%; methylome response is near-zero in both groups). In these representative datasets, genetic nomination identified features that responded no more than the rest, not features the interventions failed to move. Scales are layer-specific (proteome/metabolome in SD units, transcriptome log2 fold-change, methylome |Δβ|) and not comparable across panels. Source data: source_data/perfeature_magnitude.tsv.

### Apparent methylation persistence is sensitive to baseline variance

The methylome was the pre-specified test of the hypothesis that genetically nominated marks resist reversal — the epigenetic “memory” reported in adipose tissue [8]. In raw data, nominated CKM CpGs appeared to persist. In strong-perturbation bariatric adipose data they responded less than comparison features (Benton; permutation *P* = 0.020–0.021 at the two nomination tiers). This persistence was sensitive to baseline variance. Nominated CKM CpGs occupied low-variance regions of the methylome. They were enriched in CpG islands (odds ratio ≈ 1.20) and promoters (odds ratio ≈ 1.31, *P* = 2 × 10⁻⁸). They also had lower inter-individual baseline standard deviation than comparison features in all five datasets (Mann–Whitney *P* = 1.7 × 10⁻⁶ to 4 × 10⁻³ at the primary tiers; Supplementary Table S6a). A feature with a smaller baseline dynamic range can change less under any perturbation. Consistent with a compositional explanation, within CpG islands nominated and comparison CpGs had indistinguishable baseline variance (Mann–Whitney *P* = 0.24). This non-significant within-stratum comparison is compatible with, but does not by itself prove, equality.

Matching comparison features on baseline variance attenuated the apparent persistence to null. Variance-matching attenuated every methylome contrast to non-significance at the tier reported in Table 2 (Benton, permutation *P* = 0.57–0.76 across nomination tiers; DIRECT-PLUS, the largest paired methylation cohort at n = 256, *P* = 0.065 at the SMR-primary tier). Three independent corrections did the same: ordinary-least-squares adjustment for baseline variance, restriction to a narrow baseline-SD band, and M-value rescaling. The cell nearest to off-null was the variance-matched DIRECT-PLUS cell at the SMR-primary tier (Cliff’s δ = +0.031, 95% CI −0.003 to +0.064; Fig. 2), in the *respond-more* direction; its label-permutation test is non-significant (variance-matched *P* = 0.065, interquartile range 0.036 to 0.098 over 40 matched draws). Both the effect size and the permutation *P* are medians over 40 independent variance-matched draws, because a single draw is not a property of the data: the point estimate is positive in all 40 draws, but the interval that propagates the draw includes zero, where any single draw can exclude it (Fig. 4). Across the 15 cohort-by-tier cells, 14 of 15 variance-matched contrasts did not reach permutation significance; the exception is DIRECT-PLUS at the broader MR-significant tier (*P* = 0.045, interquartile range 0.028 to 0.075), whose interquartile range straddles 0.05 (Fig. 4d; Supplementary Table S6a; Table 2 displays one tier per cohort). Taken together, these analyses provide no consistent evidence of a material difference between nominated and comparison features. Because functionally constrained CpGs are systematically lower-variance, comparisons between feature sets that differ in baseline variance on methylation data may recover an apparent persistence that variance-matching removes. Baseline variance should therefore be assessed in such analyses.

**Figure 4.**
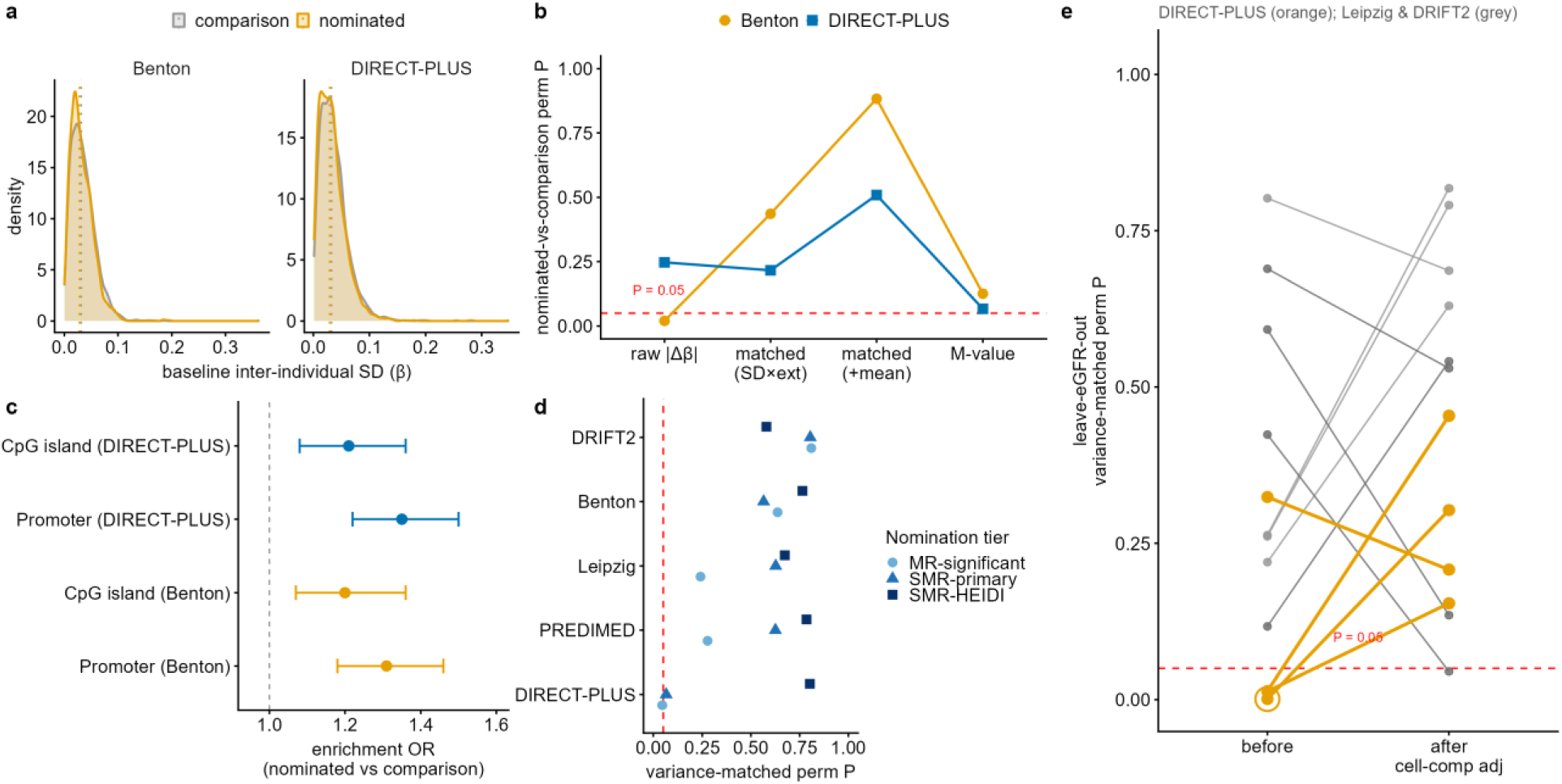
An apparent methylation persistence is sensitive to baseline variance. **(a)** Baseline inter-individual standard-deviation (β) distributions for nominated versus comparison CpGs in two representative cohorts — the bariatric-surgery Benton cohort and the Mediterranean-diet DIRECT-PLUS cohort: nominated CKM CpGs are shifted toward lower variance. **(b)** The apparent persistence attenuates to non-significance on adjustment: relative to the raw |Δβ| contrast, the nominated-versus-comparison permutation *P* is non-significant under variance-matching (on baseline SD and mean-extremity, and again under a separate match on baseline SD and baseline mean) and under M-value rescaling — every adjusted *P* exceeds 0.05 in both cohorts. The attenuation is in Benton, whose raw contrast was the significant one; DIRECT-PLUS’s raw contrast was already non-significant. **(c)** Nominated CpGs are enriched in CpG islands and promoters (enrichment odds ratio > 1 in both DIRECT-PLUS and Benton). **(d)** Matched permutation *P* across all cells: every methylation cohort (DIRECT-PLUS, PREDIMED, Leipzig, Benton, DRIFT2) × nomination tier (MR-significant, SMR-primary, SMR-HEIDI). Fourteen of the fifteen exceed 0.05; the exception is DIRECT-PLUS at the MR-significant tier (0.045), whose interquartile range across matched draws straddles the threshold. **(e)** Cell-composition adjustment removes the single-cohort signal: in the three blood cohorts under the leave-eGFR-out nomination, the variance-matched permutation *P* before versus after adjusting each CpG’s within-person change for the within-person change in blood cell composition (EpiDISH). DIRECT-PLUS’s single-cohort signal (the blip; broadest-tier *P* = 1 × 10⁻³) rises above 0.05 after adjustment, while Leipzig and DRIFT2 remain non-significant at every tier after false-discovery control (one Leipzig cell reaches *P* = 0.045 before correction; minimum *q* = 0.54) — consistent with influence from the leukocyte-proportion shift accompanying weight loss rather than a property specific to nominated CpGs. Source data: source_data/{methylation_contrast_summary.tsv, variance_dist_sample.csv}; cell-composition contrast in Supplementary Table S10a.

Two further checks supported that the methylome result is not driven by estimated-glomerular-filtration-rate power or by blood cell composition, and we report both. Because estimated glomerular filtration rate dominates the nominated CpG set, we repeated the variance-matched contrast excluding eGFR-nominated CpGs (leave-eGFR-out). It remained non-significant in the two other blood cohorts (Leipzig and DRIFT2; no nomination tier reached significance after matching). But in the largest cohort, DIRECT-PLUS, nominated CpGs appeared to respond *more* than comparison features (matched *P* = 1 × 10⁻³ at the broadest tier). This single-cohort signal did not replicate. It attenuated to non-significance after adjusting each CpG’s within-person change for the within-person change in blood cell composition, estimated by reference-based deconvolution (EpiDISH; DIRECT-PLUS matched *P* rose to 0.21–0.45). This is consistent with, but does not by itself establish, influence from the shift in leukocyte proportions that accompanies weight loss (Supplementary Table S10b) rather than a property specific to nominated CpGs. Under the eGFR-inclusive analysis the contrast was null both before and after cell-composition adjustment in all three blood cohorts, with no nomination tier significant after false-discovery control (Supplementary Table S10a). The two nominated-set definitions therefore diverge in only one cohort and for this resolvable reason.

### The result is stable across interventions, platforms and nomination tiers

The result was stable across interventions, platforms and nomination-stringency tiers. The 13 magnitude analyses span diet, bariatric surgery, sodium-glucose cotransporter-2 inhibition (empagliflozin, EMPEROR; permutation *P* = 0.72) and behavioral weight loss. Two further interventions — semaglutide (glucagon-like-peptide-1 receptor agonism) and exercise training — were examined as response-enrichment checks rather than as magnitude cells. Neither reached significance for preferential responsiveness (exercise: 61% of nominated versus 49% of comparison proteins moved restoratively, OR 1.64, 95% CI 0.79–3.40, *P* = 0.18). It held across two proteomic platforms whose nominations were independently derived and only partially overlapping, Olink/UKB-PPP and SomaScan/deCODE. These converged on seven cross-platform nominated proteins (TNFSF12, APOH, ARG1, MMP12, LRIG1, IDI2, SPON1; Supplementary Table S8a), none of which responded preferentially. Empagliflozin, a kidney-protective drug, did not show evidence of preferential response among the proteins nominated for kidney and heart-failure outcomes (n = 143; permutation *P* = 0.99).

Two factors that could have biased the comparison toward a null did not materially change the conclusion. First, the nominated sets were dominated by estimated glomerular filtration rate through GWAS power. But removing eGFR (leave-eGFR-out) and capping each outcome’s contribution (per-outcome-balanced) did not change the conclusion in the proteome, transcriptome or metabolome. The one leave-eGFR-out signal that emerged — in the largest methylation cohort — did not replicate and was consistent with influence from blood cell composition (above). The conclusion was therefore not an artifact of a single outcome. Second, metabolite nominations are inflated by pleiotropy and by reverse causation (serum metabolites are themselves filtered by the kidney). Yet restricting the nominated metabolite set to those surviving weighted-median and MR-Egger pleiotropy filters left the result unchanged (permutation *P* = 0.38 and 0.25). Across all sensitivity cuts examined, genetic nomination was not associated with larger response magnitude.

The null also held across the dependence and instrument structure of the data (Fig. 5). It held in every nomination-stringency tier of the proteome, transcriptome and metabolome, and in 14 of the 15 methylome cohort-by-tier cells (the exception, DIRECT-PLUS at the MR-significant tier, *P* = 0.045, is discussed above). The cis-instruments were uniformly strong (100% with *F* > 10; median *F* = 367 and 85 for the proteome and metabolome; Supplementary Table S4). 72% of proteome cis-loci carry multiple conditional signals, so single-variant colocalization can be incomplete. Even so, the Mendelian-randomization-tier analyses, which require no colocalization, showed no evidence of preferential response either. Clustering metabolites by their shared metaboQTL gene (690 metabolites collapsing to 454 independent loci) widened the confidence intervals only modestly and left every contrast null. The one systematic difference between the groups was that nominated proteins were better-instrumented than comparison proteins (median cis-instrument *F* = 800 versus 373; Mann–Whitney *P* < 0.001; Supplementary Table S6b). This is an expected consequence of requiring features to pass Mendelian randomization. Instrument strength is a property of the genetic architecture rather than of the response assay and does not directly bear on response magnitude. We cannot, however, fully exclude residual differences between the groups in response-assay baseline mean, variance or detection, which remains a limitation.

**Figure 5.**
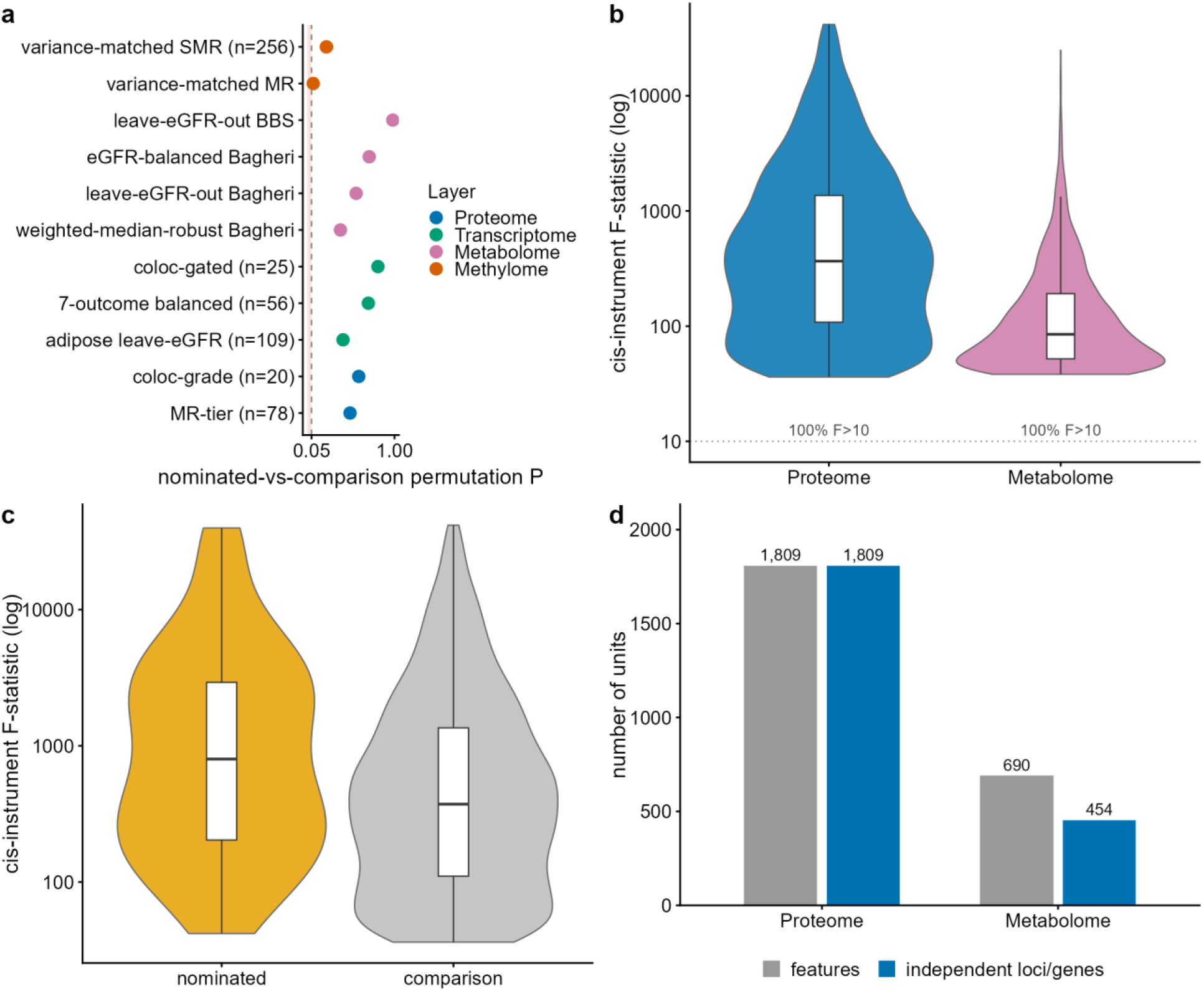
Sensitivity of the null to nomination stringency, instrument strength, baseline balance, and feature dependence. **(a)** Nomination-stringency robustness: nominated-versus-comparison permutation *P* for every stringency tier × layer; all exceed 0.05 except the methylome’s MR-significant tier (*P* = 0.045, interquartile range 0.028 to 0.075 across matched draws; panel d of Fig. 4), so the result did not vary materially with the nomination threshold. **(b)** Instrument strength: cis-instrument F-statistic distributions (log) for the proteome and metabolome; 100% exceed F = 10 (median 367 and 85). 72% of proteome cis-loci are multi-signal, so conventional single-variant colocalization can be incomplete — but the MR-tier analyses, which require no colocalization, are also null. **(c)** Baseline-property balance: cis-instrument F-statistic for nominated versus comparison proteins (leave-eGFR-out set); nominated proteins are better-instrumented (median 800 versus 373; Mann–Whitney *P* < 0.001), an expected and disclosed imbalance that does not bear on response magnitude. **(d)** Dependence-aware resampling unit: 690 metabolites collapse to 454 distinct metaboQTL genes (0.66×), so cluster-aware intervals are modestly wider but the contrasts remain null; the proteome is already locus-level (each protein = one cis gene; the 1,809 cis-instrumented proteins are the resampling universe, of which 1,607 are also on the DiRECT SomaScan panel and enter the representative contrast as 1,736 aptamer measurements — 85 nominated + 1,651 comparison, shown in Fig. 1b). Source data: source_data/stringency_robustness.csv (methylome rows derived at render time from source_data/methylation_contrast_summary.tsv), source_data/rob_*.tsv.

### Cross-layer concordance is limited by mapping and sample size

If response were targeted at nominated biology, the multiple molecular features of a gene nominated in more than one layer might respond together. Within the available data they did not separate from chance (Supplementary Fig. S2). Among 74 genes nominated in at least two layers (Supplementary Table S8b), the features responded in a concordant health-protective direction in 45 of 84 layer-pairs (54%, 95% CI 43–64%). This was indistinguishable from the concordance expected under independent response by a within-node permutation that preserves each layer’s marginal restorative rate (permutation *P* = 0.29). No gene was nominated in all four layers, and three-layer overlap was limited to five genes: three shared by the methylome, proteome and metabolome (DPEP1, GATM, GCKR) and two by the proteome, transcriptome and methylome (NFATC1, SORT1). The test is therefore necessarily of pairwise coherence. The analysis is underpowered. With 84 pairs it cannot exclude concordance up to ∼64%, and we present it as the limit of what current public data support, not as a positive null. Bridging genes across layers by symbol, and mapping metabolites to genes through the nearest metaboQTL gene, is itself approximate, further limiting node-level inference.

One stratum is hypothesis-generating but not robust. Layer-pairs that excluded the methylome were 76% concordant (13 of 17; *P* = 0.05), driven by metabolite–protein enzyme nodes (8 of 10), whereas pairs involving the methylome were at chance (48%). This split is partly biological and partly structural. Methylation response magnitudes are near zero (median |Δβ| = 0.0013), so restorative-direction calls in the methylome are correspondingly unstable and cannot contribute coherence by construction. We carry the metabolite–protein coherence as a hypothesis for a powered test, not as a finding, and exclude it from our conclusions.

### A secondary, exploratory directional analysis (inconclusive)

On the secondary axis — whether nominated features moved opposite to their disease-associated Mendelian-randomization direction — a sub-threshold asymmetry appeared in the proteome. It is reported in full in the Supplement (Supplementary Fig. S4, Supplementary Table S7). Among colocalization-grade nominated Olink proteins, 70% moved opposite the disease direction versus 49% of comparison features, with a concordant pattern on SomaScan (57% versus 47%). The asymmetry did not survive correction. Against a margin-preserving permutation null that shuffles disease-associated signs among nominated features (the appropriate null when sign margins are imbalanced), the SomaScan fraction was non-significant (*P* = 0.12). The Olink fraction was also non-significant on a one-group test (*P* = 0.12). The effect did not replicate in the transcriptome, methylome or metabolome, and no directional test survived false-discovery control within the directional family. The 0.5 binomial null is not generally valid, and the most direct test of restoration — within-person response against incident events — cannot be performed in summary-level data. We therefore treat the directional axis as inconclusive and do not interpret it as evidence of clinical benefit.

## Discussion

Across the four molecular layers and intervention datasets examined, features meeting the selected genetic-nomination criteria did not show a consistent tendency toward larger molecular-response magnitudes than adequately-instrumented comparison features. This is a comparative result, not evidence that nominated features were unchanged. Many nominated proteins and metabolites changed substantially after intervention, and the contrast estimates the relative distribution of response, not whether any individual feature was engaged. The data therefore indicate limited concordance between these genetic-evidence categories and short-term response magnitude, while the smaller proteomic and transcriptomic analyses remain compatible with modest differences. “Non-nominated” does not mean non-causal. A feature can be genuinely disease-relevant yet respond no more than its comparators.

The methylation analyses identify a measurement issue likely to generalize. Nominated CpGs occupied genomic contexts — CpG islands and promoters — with lower baseline inter-individual variance. This made their raw absolute-change distributions difficult to compare with those of the broader comparison set. The apparent difference attenuated after matching on baseline variance and related sensitivity analyses. This is a narrower test than Hinte et al.’s [8], which demonstrated persistent transcriptional changes in human and mouse adipose and persistent epigenomic changes in mouse adipocytes, and it does not refute epigenetic memory. Rather, it shows that baseline dynamic range and genomic context should be evaluated whenever molecular change or persistence is compared across selected feature sets. This extends, in a measurement sense, the single-layer observation that disease-associated transcripts are frequently disease-induced rather than disease-causing [9].

Several limitations constrain interpretation, and one is central. Genetic nomination is power-dependent, the “nominated if any outcome” union favors better-powered outcomes, and false-negative nominations would dilute the group contrast toward zero. Non-nomination therefore does not establish absence of disease relevance, and the comparator may contain disease-relevant features. Nomination criteria differed across layers. Colocalization, SMR/HEIDI and metabolite pleiotropy filters have different error structures and are combined only as a descriptive cross-layer framework, not a uniform causal classifier. Response cohorts differed in tissue, intervention, estimand and sample size (median 118 participants, range 15 to 1,134), and molecular features within a layer are correlated. Our confidence intervals are computed at the feature level and do not capture participant-level sampling uncertainty. They should therefore be read as descriptive rather than as precise bounds, and we do not draw a formal equivalence conclusion. Cross-layer gene and metabolite-to-gene mapping is approximate. These limitations preclude conclusions about biological orthogonality of causality and reversibility, residual-risk mechanisms, or therapeutic targets.

These observations bear on how genetic target evidence is used in translational practice, where support from human genetics is associated with greater success in clinical development. Genetic evidence is increasingly invoked not only to prioritize targets but also, in some proposals, to choose which molecular features to track as treatment-response readouts and to define strata of patients expected to respond. That evidence indexes a feature’s lifelong, germline-anchored association with disease — a different quantity from how much its molecular readout moves over the weeks-to-months horizon of an intervention. Our comparative results are compatible with limited concordance between these two properties. Within the layers, cohorts and nomination definitions studied, features carrying stronger CKM genetic support were not consistently associated with larger short-term molecular responses. A practical reading is that target-nomination genetics remains well suited to selecting and de-risking targets, but on these data should not by itself be repurposed as a response or reversibility biomarker. Such repurposing would mean choosing among candidates on expected responsiveness, selecting molecular markers to monitor therapy, or stratifying patients by anticipated molecular benefit. This caution is bounded rather than absolute. Non-nomination does not mark a feature as unresponsive or as lacking disease relevance. The smaller proteomic and transcriptomic analyses remain compatible with modest differences that a response-informative use case would need to establish prospectively. Response-biomarker and stratification candidates therefore continue to warrant selection and qualification on measured response and outcome in their intended clinical context. Genetic nomination is one descriptive input among several rather than a stand-in for that evidence.

The decisive test is therefore a different design. Standardized, same-subject longitudinal studies that measure intervention exposure, molecular change and incident CKM events are needed to determine whether molecular change in genetically nominated features carries prognostic or therapeutic information beyond the interventions themselves. Such studies would incorporate matched or continuous genetic-evidence models, participant- and feature-cluster-aware inference, and non-European ancestries. The present comparative result and the methylation-variance lesson motivate, but cannot substitute for, that experiment.

In sum, across four molecular layers and a range of weight-loss and cardiometabolic interventions, genetic nomination for CKM association did not consistently identify features with systematically larger molecular responses. Apparent methylation persistence was sensitive to baseline variance. Whether molecular change in genetically nominated features carries prognostic or therapeutic information beyond the interventions themselves will require same-subject longitudinal studies linking within-person molecular change to incident CKM outcomes.

## Methods

### Study design

We tested, in each of four molecular layers, whether features genetically nominated for CKM association respond to weight-loss and cardiometabolic interventions differently from non-nominated comparison features. The design is genetics-anchored and two-sample. Nomination status was assigned from germline cis-QTL Mendelian randomization against disease GWAS, and molecular response was measured in independent paired human intervention cohorts. The nominated-versus-comparison response-magnitude contrast was the pre-specified primary endpoint. Restorative direction and cross-layer node concordance were secondary. No prior hit lists were imported. Every adequately-instrumented feature in each response dataset was classified de novo. Only adequately-instrumented features entered the comparison group, to reduce differential opportunity for nomination arising solely from the absence of an instrument. Residual differences in QTL coverage, tissue relevance and GWAS power remain.

### Genetic nomination

For each layer we assembled cis-acting genetic instruments and performed Mendelian randomization against eight CKM-disease GWAS: type 2 diabetes (Xue et al. [22], GCST006867), coronary artery disease (Nikpay et al. [23], GCST003116), atrial fibrillation (Nielsen et al. [24], GCST006414), heart failure (Levin et al. [25], GCST90162626), ischemic stroke (GIGASTROKE [26], GCST90104540), chronic kidney disease, stage 3 (Million Veteran Program [36], GCST90476128), estimated glomerular filtration rate (Stanzick et al. [27], GCST90103634) and non-alcoholic fatty liver disease (Ghodsian et al. [28], GCST90091033). The outcome-GWAS discovery samples were 62,892 cases and 596,424 controls (type 2 diabetes), 60,801 and 123,504 (coronary artery disease), 60,620 and 970,216 (atrial fibrillation), 115,150 and 1,550,331 (heart failure), 62,100 and 1,234,808 (ischemic stroke), 43,088 and 394,787 (chronic kidney disease) and 8,434 and 770,180 (non-alcoholic fatty liver disease); estimated glomerular filtration rate, the sole quantitative trait, was analyzed in 1,004,040 individuals. Instruments were cis-pQTL for the proteome (UKB-PPP Olink, Sun et al. [10]; deCODE SomaScan, Ferkingstad et al. [11]), cis-eQTL for the transcriptome (GTEx liver and subcutaneous adipose, eQTL Catalogue datasets QTD000266 and QTD000116 [12,13]), cis-mQTL for the methylome (GoDMC blood [14]) and cis-metaboQTL for the metabolome (Chen et al. CLSA, independent genome-wide-significant sentinels [15]).

Instruments were harmonized to the outcome by effect allele, with strand-ambiguous palindromic SNPs at intermediate frequency (0.42–0.58) removed. Instrument strength was required to exceed F = 10. Of the three instrumental-variable assumptions, only relevance is directly testable, and it was supported by the F > 10 threshold. Independence of the instrument from confounders of the feature–outcome relationship and the exclusion restriction (the instrument acts on the outcome only through the feature) cannot be verified directly in summary-data Mendelian randomization. We sought to reduce violations by cis-restriction of instruments to the feature’s encoding locus (which limits horizontal pleiotropy) and by shared-causal-variant filtering through colocalization or SMR–HEIDI. We added weighted-median, MR-Egger and Steiger-directionality checks in the pleiotropy-prone metabolome.

We estimated the per-feature Mendelian-randomization effect on each outcome by the Wald ratio for single-instrument features and by inverse-variance-weighted regression for multi-instrument features. We controlled the false-discovery rate within each outcome (Benjamini– Hochberg [38] q < 0.10); the resulting per-feature adjusted q-value (the minimum across the outcomes for which a feature was nominated) is reported for every nominated feature in Supplementary Table S2.

High-confidence nomination was assigned by colocalization (coloc.abf [29]; posterior probability of a shared causal variant PP.H4 ≥ 0.8 and PP.H4/PP.H3 ≥ 5) for the proteome and transcriptome. For the methylome it was assigned by summary-data-based MR with the HEIDI heterogeneity test [30]. SMR with HEIDI supports an association consistent with a shared causal variant under its assumptions and is not by itself evidence that the feature mediates disease. We therefore describe methylome calls as genetically-supported shared associations. Conventional coloc.abf assumes a single causal variant per region, which can reduce accuracy at multi-signal loci. We treated nomination strength continuously across stringency tiers (Fig. 5) to bound this. For the metabolome, where instruments lie at pleiotropic metabolic-hub loci, we additionally computed weighted-median [32] and MR-Egger [31] estimates and applied Steiger directionality filtering [37]. A feature was labeled genetically nominated if it passed its layer’s high-confidence tier for at least one outcome, and non-nominated otherwise (Supplementary Table S2). Established disease genes were recovered as calibration examples (PCSK9, IL6R and APOB for coronary disease; GCKR for type 2 diabetes; Fig. 1c), indicating sensitivity where established causal biology exists rather than validating specificity.

Nominated-set size tracked the statistical power of the outcome GWAS. Estimated glomerular filtration rate, the only quantitative trait and the best-powered GWAS, dominated every layer. Nominated-set size varied markedly across the eight outcomes, with eGFR yielding by far the largest nominated set, consistent with differential power. We therefore pre-specified two power-robust primary analyses: leave-eGFR-out (nominated for any non-eGFR outcome) and per-outcome-balanced (the union of a fixed quota of the most-significant nominated features per outcome, so no single outcome dominates). We report eGFR-inclusive sets as secondary. This power-robust hierarchy is primary for the proteome, transcriptome and metabolome. For the methylome the headline contrast is instead the variance-matched eGFR-inclusive analysis (because baseline inter-individual variance, not outcome-GWAS power, is the dominant methylome confound), with leave-eGFR-out as a robustness check (see “Methylome baseline-variance correction”). Because nomination uses a within-outcome false-discovery rule and a “nominated if any outcome” union, we report nomination counts by outcome and layer. We treat nomination strength continuously in sensitivity analyses rather than relying only on the binary label.

### Molecular-response (intervention) data

Per-feature intervention effects came from paired human data (cohorts, assays, estimands and achieved weight change summarized in Table 1 and Supplementary Table S5a). Data were harmonized by direction only because the layers are measured on non-comparable scales. Proteome response used DiRECT [34] (caloric restriction, SomaScan) and By-Band-Sleeve (bariatric surgery, Olink) from Goudswaard et al. [4], semaglutide (STEP [16]), empagliflozin (EMPEROR, Olink [17]) and exercise training (HERITAGE [20]). Transcriptome response used liver (GSE83452 [45]; diet and bariatric, n = 79 paired) and subcutaneous adipose (GSE84046 [46]; an energy-restriction trial of 22 participants randomized to high-protein versus normal-protein arms). Methylome response used five blood and adipose EPIC/450K cohorts: DIRECT-PLUS [39] (Mediterranean diet, n = 256, the largest paired methylation cohort), the PREDIMED-Navarra methylation subset (Arpón et al. [41]; the trial itself is ref [35]), the Leipzig bariatric cohort (Müller et al. [40]), DRIFT2 [42] and Benton (RYGB adipose) [43]. Metabolome response used the peer-reviewed Bagheri/Tanriverdi cohort [44] (sleeve gastrectomy and RYGB, Metabolon) and the By-Band-Sleeve arm (Smith et al. [47]) and the DiRECT arm (Corbin et al. [21]).

The response estimand differs across datasets: within-person post-versus-baseline change in single-arm cohorts versus published treatment-versus-control effects in randomized trials. We therefore record each dataset’s estimand and present pharmacologic and behavioral interventions as separate strata. These interventions are not pooled as a single “weight loss” effect. Semaglutide and empagliflozin in particular have weight-independent pharmacologic effects (EMPEROR is a heart-failure treatment contrast). We treat them as mechanistic sensitivity strata rather than as weight-loss evidence.

### Nominated-versus-comparison contrast

The primary endpoint was the difference in molecular response magnitude between nominated and non-nominated comparison features, quantified by Cliff’s δ. Cliff’s δ is a rank-based, scale-free effect size. It is the probability that a randomly chosen nominated feature responds more than a randomly chosen comparison feature, rescaled to [−1, 1] and centered at zero under the null. Formally, δ = 2 × P(nominated > comparison) − 1, computed from the Mann–Whitney statistic. We chose it because the four layers are measured on non-comparable units (we harmonized by direction only), and because a rank statistic is robust to outliers and makes no distributional assumption. Values of |δ| = 0.10 as a small-effect reference point for descriptive orientation only; we do not treat it as a formal equivalence bound, and note that the familiar 0.1/0.3/0.5 tiers are conventions for standardized mean differences rather than stochastic dominance [18]. Confidence intervals were obtained by bootstrap resampling (1,000 replicates) and significance by label-permutation (≥ 10,000 permutations of the nominated/non-nominated labels).

Because features are not statistically independent — metabolites share metaboQTL hubs and CpGs share co-methylation blocks — feature-level intervals can be anticonservative. We therefore quantified dependence in the metabolome (the layer where clustering is greatest) by collapsing its 690 metabolites onto their 454 shared metaboQTL genes, which widened the intervals only modestly and left the contrasts null. We treat the locus/gene as the appropriate resampling unit where dependence is material. We also reported per-layer cis-instrument strength (F-statistic, proportion of multi-signal loci) and the balance of instrument strength between groups.

We synthesized the 13 analyses by DerSimonian–Laird [33] random-effects meta-analysis of the per-analysis Cliff’s δ (weighted by the inverse of the squared bootstrap standard error, reporting τ² and I²), overall and stratified by molecular layer and intervention class. A random-effects model was used because the analyses differ in measurement (platforms, tissues and scales) and so may estimate heterogeneous true effects. An I² near zero indicates between-analysis variance no larger than within-analysis sampling error. We interpret each interval descriptively rather than as a formal equivalence test. An interval whose upper bound lies below a reference rank advantage of δ = 0.10 — a conventional threshold for a small effect — is described as inconsistent with advantages larger than this. An interval spanning both negligible and modest advantages is described as inconclusive. We do not assert statistical equivalence, which would require a smallest effect of interest specified before the analysis.

The restorative (directional) endpoint asked whether the response moved a feature opposite to its disease-associated direction. The harmful direction was the sign of the feature’s Mendelian-randomization effect toward disease (inverted for eGFR, where higher values are protective). For a feature nominated for more than one outcome the direction was taken from the most-significant outcome. This rule can be vulnerable to winner’s curse and can mask sign discordance. We therefore additionally report feature–outcome-pair directions and the count of sign-discordant features in the supplement. For the exploratory directional axis, the primary test is a margin-preserving permutation. It shuffles disease-associated signs among nominated features while holding the sign margin fixed, because the 0.5 binomial null is not generally valid when disease-associated signs are imbalanced. As descriptive secondary summaries we additionally report the restorative fraction among nominated features against a null of 0.5 (by exact binomial test) and the nominated-versus-comparison odds ratio (per-layer and Mantel– Haenszel-pooled). The odds ratio additionally depends on the comparison group’s noisy sub-threshold signs. The directional axis is treated as exploratory throughout. Tests were grouped into six families (Supplementary Table S12): the 13 magnitude analyses, the directional analyses, the nomination-stringency tiers, the methylome variance and enrichment tests, the cross-layer node-concordance tests and the continuous-evidence test. False-discovery control was applied within, not across, families.

### Methylome baseline-variance correction

In the methylome, the raw nominated-versus-comparison contrast is confounded. Nominated CpGs are enriched in CpG islands and promoters and have lower inter-individual baseline standard deviation than comparison CpGs, which compresses their response range. We therefore pre-specified variance-matching as the primary methylome contrast. Comparison CpGs were subsampled to match the nominated set on two baseline properties before the permutation test: the per-CpG inter-individual standard deviation of β, and mean-extremity, defined as the absolute deviation of the baseline mean β from 0.5 (a CpG close to 0 or 1 has less room to move than one near 0.5). Both variables were converted to ranks and cut into quantile strata over the combined nominated-plus-comparison universe — 20 standard-deviation strata by 10 extremity strata, 200 cells — and within each cell up to five comparison CpGs were drawn per nominated CpG, without replacement and from a fixed seed. A cell holding fewer than five per nominated CpG contributed all of them, and a cell with no comparison CpG contributed none; the nominated arm was never trimmed, so the matched comparison set is at or just below five times the nominated count. Because the matched permutation *P* proved sensitive to which comparison CpGs were drawn — 40 independent draws of one cell spanned *P* = 0.001 to 0.196 — the reported value is the median over 40 draws, with the interquartile range given beside it; a single draw is not a property of the data. This is the contrast labeled “matched (SD × ext)” in the methylome adjustment panel. The neighboring “matched (+ mean)” bar is a separate hardening analysis rather than a third matching variable: it re-matches on 12 baseline-standard-deviation by 12 baseline-mean strata and additionally adjusts for baseline mean in the ordinary-least-squares model. The confound and its attenuation were examined by four independent approaches: ordinary-least-squares adjustment of |Δβ| for baseline variance, restriction to a narrow baseline-SD band, M-value rescaling, and a CpG-island composition analysis (testing baseline variance of nominated versus comparison CpGs within islands). EPIC/450K IDATs were processed with minfi (Noob normalization). As a post-hoc sensitivity restricted to the blood cohorts (DIRECT-PLUS, Leipzig, DRIFT2), blood cell-type composition was estimated by reference-based deconvolution (EpiDISH robust partial correlations, centDHSbloodDMC reference). The nominated-versus-comparison contrast was repeated after adjusting each CpG’s within-person change for the corresponding within-person change in estimated cell-type proportions. No tier was significant after false-discovery control (Supplementary Table S10a). The per-CpG adjustment is interpretable only where the paired sample size supports it (it is degrees-of-freedom-limited in the small cohorts). Adipose data (Benton) were not deconvolved with a blood reference. Because the methylome’s dominant confound is baseline inter-individual variance rather than outcome GWAS power, its headline contrast is the variance-matched eGFR-inclusive analysis. We additionally report the leave-eGFR-out nominated set and a blood cell-composition adjustment, each before and after, as power- and composition-robustness checks (Supplementary Table S10a). Because baseline mean, variance, assay reliability and detection rate can differ between nominated and comparison features in any layer, we additionally examined baseline-property balance in the proteome, transcriptome and metabolome.

### Cross-layer node concordance

For genes nominated in at least two layers, each layer’s feature was reduced to a restorative call (responds toward health, yes/no) using the harmful direction defined above. Cross-layer concordance was the fraction of layer-pairs, within a node, that agreed in their restorative call. The 84 layer-pairs are nested within 74 genes, and chance agreement depends on each layer’s marginal restorative rate (it is p₁p₂ + (1 − p₁)(1 − p₂), not automatically 0.50). We therefore tested the observed concordance against the permutation-expected concordance using a within-node permutation that preserves each layer’s marginal restorative rate. We report the permutation-expected value and its interval rather than a fixed 50% line. Genes were bridged across layers by symbol. Metabolites were mapped to genes through the nearest protein-coding gene of their metaboQTL and CpGs through annotated gene region — both approximate assignments that limit node-level inference. Because the methylome’s response magnitudes are near zero (median |Δβ| = 0.0013), its restorative direction is largely uninformative, and we report methylome-stratified results.

### Statistics and Reproducibility

No statistical method was used to predetermine sample size. This is a secondary reanalysis of existing datasets. The intervention-cohort sizes (median 118 participants, range 15 to 1,134; Table 1) were fixed by the source studies and could not be altered. The number of features per layer was set by the assay platforms and cis-QTL atlases rather than by a power calculation. Data inclusion and exclusion followed the pre-specified adequately-instrumented rule. Only features with a cis-instrument passing the F > 10 relevance threshold entered the comparison, and strand-ambiguous palindromic variants at intermediate frequency (0.42–0.58) were removed during harmonization. No ad hoc exclusions were applied to the final feature-, cohort- or data-point inclusion rules. The investigators did not randomize samples and were not blinded to nomination status. The analysis reuses previously randomized or observational datasets and is deterministic and analytic. Nomination status and molecular response are each derived from published summary or per-feature statistics, so experimental randomization and blinding are not applicable. The core statistical approach is detailed in “Nominated-versus-comparison contrast” above: rank-based Cliff’s δ as the primary effect size, bootstrap confidence intervals interpreted descriptively rather than as precise exclusion bounds, label-permutation for significance, Benjamini–Hochberg false-discovery control applied within (not across) test families, and DerSimonian–Laird random-effects synthesis of the per-analysis estimates.

Mendelian-randomization and contrast engines were implemented in Python. Colocalization (coloc), summary-data-based MR (SMR/HEIDI), methylation processing (minfi) and figures (ggplot2) were implemented in R. Software and package versions, random seeds and the frozen analysis scripts are provided (Supplementary Table S9). The analysis code and frozen source data for every figure and table are available at https://github.com/badboybert/ckm-4layer-null and archived at Zenodo (concept DOI 10.5281/zenodo.20965271; version used here, v1.2.6). The study was not prospectively registered. The primary, secondary and post-hoc status of each analysis is declared in the master analysis specification (Supplementary Table S11, Additional file 2). Headline analyses were independently re-derived from the raw input files by a separate pipeline run from the frozen scripts against pre-specified numerical acceptance criteria. Discrepancies were reconciled to the canonical engines, and analyses that failed re-derivation — for example an early restorative-direction comparator that compared against an inappropriate baseline set — were corrected or removed before the results reported here. We report instrument and Mendelian-randomization details following the STROBE-MR guideline [19]. Any use of generative-AI assistance in analysis or drafting is disclosed in accordance with the journal’s policy.

### Ethics and data-reliance statement

This study is a secondary reanalysis of previously collected, publicly available or consortium-released data. These comprise genome-wide association and cis-QTL (pQTL, eQTL, mQTL, metaboQTL) summary statistics, and molecular-response data from prior intervention studies. The molecular-response data comprise per-feature intervention effect estimates reported in prior publications together with de-identified individual-level molecular measurements (for example per-sample DNA-methylation and gene-expression matrices) drawn from open-access repositories (GEO and ArrayExpress) under the original studies’ terms of use (Table 1; References). We recruited no human participants, generated no new primary data, and accessed no individual-level identifiable participant records. Each contributing dataset was generated under the ethics approval and participant informed consent reported in its primary publication, to which we refer readers for those details. This work is a secondary analysis of de-identified data that were already collected and released by the original studies. It involves no new human-participant recruitment and no access to identifiable individual-level records. No additional ethics approval was required for the present analysis. Every dataset was obtained either from an open-access repository or from a published article’s supplementary tables; we applied for no controlled-access individual-level data, so no controlled-access application or data-use agreement was required for this analysis. The study was conducted in accordance with the principles of the Declaration of Helsinki.

### Inclusion and multi-ancestry statement

We recruited no participants and generated no new primary data. Questions of local-researcher inclusion in the primary-data sense therefore do not apply to this secondary analysis. The genetic instruments and most contributing intervention cohorts are of predominantly European ancestry (Table 1; Supplementary Tables S5a and S5b), which we note as a limitation on generalizability. The eight outcome genome-wide association studies are likewise predominantly European: 95% of their discovery participants. Two are materially multi-ancestry — coronary artery disease (25% non-European) and heart failure (18% non-European) — while for stroke, kidney function and chronic kidney disease we analyzed the European-ancestry stratum released by studies that also published non-European strata, so the ancestral diversity of those parent studies does not extend to the data analyzed here. The nominated-versus-comparison contrasts are estimated within, and their transportability across ancestries is not established by, the datasets studied. Replication in non-European-ancestry cohorts is needed, consistent with the same-subject, multi-ancestry longitudinal designs called for in the Discussion.

## Declarations

### CKM outcome GWAS summary statistics

(EBI GWAS Catalog accessions): type 2 diabetes, GCST006867; coronary artery disease, GCST003116; atrial fibrillation, GCST006414; heart failure, GCST90162626; stroke, GCST90104540; chronic kidney disease (Million Veteran Program), GCST90476128; estimated glomerular filtration rate, GCST90103634; non-alcoholic fatty liver disease, GCST90091033. All are available from the GWAS Catalog [48] (https://www.ebi.ac.uk/gwas/).

### Cis-QTL atlases used for genetic nomination

- *Proteome (cis-pQTL):* UKB-PPP plasma proteomics (Olink), available via the UK Biobank Pharma Proteomics Project [10] (Sun et al. 2023; http://ukb-ppp.gwas.eu/); deCODE plasma proteomics [11] (SomaScan; Ferkingstad et al. 2021; https://www.decode.com/summarydata/).
- *Transcriptome (cis-eQTL):* GTEx v8 [12] (https://gtexportal.org/) and the eQTL Catalogue [13], specifically liver dataset QTD000266 and subcutaneous adipose dataset QTD000116 (https://www.ebi.ac.uk/eqtl/).
- *Methylome (cis-mQTL):* GoDMC blood DNA-methylation QTL atlas [14] (http://mqtldb.godmc.org.uk/).
- *Metabolome (cis-metaboQTL):* Chen et al. CLSA plasma-metabolome GWAS (genome-wide-significant sentinels; Chen et al. 2023, *Nat Genet*; data in the study’s supplementary materials).

### Molecular-response (intervention) cohorts

- *Transcriptome:* Gene Expression Omnibus [49] accessions GSE83452 (liver; diet and bariatric surgery) and GSE84046 (subcutaneous adipose; energy-restriction high-protein vs normal-protein trial) (https://www.ncbi.nlm.nih.gov/geo/).
- *Proteome response:* per-protein DiRECT (SomaScan) and By-Band-Sleeve (Olink) reversal effects from the supplementary tables of Goudswaard et al. 2023 (*Sci Rep*; DOI 10.1038/s41598-023-47030-x); semaglutide (STEP), empagliflozin (EMPEROR) and exercise-training (HERITAGE) effects from the respective publications’ supplementary materials.
- *Metabolome response:* per-metabolite reversal effects from the published supplementary tables of the Bagheri/Tanriverdi sleeve-gastrectomy/RYGB cohort (Bagheri et al. 2024, *Metabolism*), of Corbin et al. 2024 (*Diabetologia*) for the DiRECT arm, and of Smith et al. 2025 (*Wellcome Open Res*) for the By-Band-Sleeve arm.
- *Methylome response:* per-CpG reversal effects from DIRECT-PLUS, PREDIMED, the Leipzig bariatric cohort (Müller et al. 2024, *EBioMedicine*), DRIFT2 and Benton (RYGB adipose), obtained from each study’s open-access repository deposit — ArrayExpress E-MTAB-3052 (Benton), E-MTAB-12527 (DIRECT-PLUS) and E-MTAB-13564 (Leipzig); GEO GSE107205 (PREDIMED-Navarra) and GSE240184 (DRIFT2).

Every dataset analyzed here was obtained either from an open-access repository or from a published article’s supplementary tables, and each is cited at first use in the Methods and reference list. Several of the per-feature effect sets we reused were published as supplementary tables by studies whose underlying individual-level data are themselves managed-access, including the By-Band-Sleeve and DiRECT proteome and metabolome sets; we used only the published summary tables and applied for no controlled-access individual-level data, so this analysis required no data-use agreement or controlled-access application.

### Frozen source data for this manuscript

The exact per-figure and per-table source data underlying all displayed results — including magnitude_effects.csv, methylation_contrast_summary.tsv, the per-layer contrast result files and the node-concordance tables — are provided as read-only snapshots in the source_data/ directory of the project repository (below) and are archived at Zenodo (concept DOI 10.5281/zenodo.20965271). Provenance for each frozen file (canonical result, generating script and the figure/table it feeds) is documented in source_data/PROVENANCE.md and the source-data manifest.

### Code availability

All analysis code — the Mendelian-randomization and contrast engines (Python), and the colocalization (coloc), SMR/HEIDI, methylation processing (minfi, EpiDISH) and figure-generation (ggplot2) scripts (R) — together with software and package versions, random seeds and the frozen analysis scripts, is available at https://github.com/badboybert/ckm-4layer-null and archived at Zenodo (concept DOI 10.5281/zenodo.20965271, which resolves to the latest version). The version used for this manuscript is tagged v1.2.6. The repository includes the frozen source_data/ snapshots and the re-derivation pipeline used to independently reproduce the headline analyses against pre-specified numerical acceptance criteria.

### Use of generative AI

The authors used generative-AI assistance (a large-language-model coding and writing assistant) to support code scaffolding, literature look-up and manuscript drafting and editing. All AI-assisted outputs — including code, statistical results and text — were reviewed, verified and edited by the authors, who take full responsibility for the integrity, accuracy and content of the work. Generative AI was not used to generate, fabricate or alter data, results or figures, and is not listed as an author. This disclosure is made in accordance with the journal’s policy on the use of AI tools.

## Author contributions (CRediT)

C.-Y.H. — Formal analysis, Investigation, Data curation, Software, Visualization, Writing – original draft. K.-T.C. — Methodology, Validation, Resources, Writing – review & editing. B.C.-M.T. — Conceptualization, Methodology, Supervision, Funding acquisition, Project administration, Writing – review & editing. All authors read and approved the final manuscript.

## Data Availability

This study is a secondary analysis of publicly available and consortium summary-level data; no new primary human data were generated. All genetic-nomination and molecular-response inputs are openly available at the sources and accessions listed in the manuscript's Data-availability section (GWAS Catalog; UKB-PPP; deCODE; GTEx; eQTL Catalogue QTD000266/QTD000116; GoDMC; GEO GSE83452, GSE84046, GSE107205, GSE240184; ArrayExpress E-MTAB-3052, E-MTAB-12527, E-MTAB-13564; and the cited studies' published supplementary materials). All derived per-figure/per-table source data and analysis code are available at https://github.com/badboybert/ckm-4layer-null and archived at Zenodo (https://doi.org/10.5281/zenodo.20965271).

## Acknowledgements

We thank the participants and investigators of the original cohorts and consortia whose summary data were used — the UK Biobank Pharma Proteomics Project, deCODE genetics, GTEx, the eQTL Catalogue, GoDMC, the GWAS Catalog and the contributing CKM outcome-GWAS consortia (Million Veteran Program, GIGASTROKE, and the heart-failure meta-analysis of Levin et al. and its contributing studies, among others), and the DiRECT, By-Band-Sleeve, DIRECT-PLUS, PREDIMED, STEP, EMPEROR, HERITAGE, Bagheri/Tanriverdi and Leipzig study teams.

**Supplementary figures. S1**, random-effects subgroup synthesis — pooled Cliff’s δ overall and by molecular layer, all within the small-effect reference region (±0.10). **S2**, cross-layer node concordance — 45/84 = 54% of layer-pairs concordant, indistinguishable from the permutation expectation and underpowered; hypothesis-generating only. **S3**, eGFR-set dominance — eGFR dominates nominated-set size, motivating the pre-specified leave-eGFR-out primary analysis. **S4**, exploratory directional analysis — restorative-direction forest (per-layer and pooled odds ratios; one interval marginally excludes 1, PREDIMED at odds ratio 0.90, 95% CI 0.82 to 0.998, and no stratum is significant after false-discovery control, minimum *q* = 0.41); the nominated-only margin-preserving permutation results are reported in the Results and Methods; inconclusive (Supplementary Table S7). Source files: source_data/{node_reversal_concordance.tsv, egfr_dominance.csv}, and Supplementary Table S7 (S7_directional.tsv).

