## Supplementary Information and Figures for "Genetically nominated cardiovascular-kidney-metabolic features are not consistently more responsive to weight-loss and cardiometabolic interventions"

### **Supplementary Tables**

**Supplementary Table S1. Feature flow per layer.** Per molecular layer: measured features, the analysis universe (the set entering each contrast, i.e. nominated + comparison, and therefore smaller than the cis-instrumented universe), the genetically nominated set, and the non-nominated comparison set. Proteome counts below the measured row are SomaScan aptamers, the unit the contrast was run on (1,736 aptamers over 1,607 distinct proteins); the methylome comparison count is the baseline-variance-matched subsample, five per nominated CpG, not the full non-nominated population, so the methylome analysis universe is much smaller than its instrumented, measured set. These are the per-representative-contrast sets (the methylome row uses the DIRECT-PLUS cohort; per-cohort methylome counts are in Table 2), a different denominator from the deduplicated nominated master list in Table S2 — the two are not directly comparable feature-by-feature.

| **layer** | **stage** | **n** |
| --- | --- | --- |
| Proteome | measured | 4170 |
| Proteome | analysis universe | 1736 |
| Proteome | nominated | 85 |
| Proteome | comparison | 1651 |
| Transcriptome | measured | 28541 |
| Transcriptome | analysis universe | 1350 |
| Transcriptome | nominated | 25 |
| Transcriptome | comparison | 1325 |
| Metabolome | measured | 952 |
| Metabolome | analysis universe | 555 |
| Metabolome | nominated | 255 |
| Metabolome | comparison | 300 |
| Methylome | measured | 866091 |
| Methylome | analysis universe | 9181 |
| Methylome | nominated | 1531 |
| Methylome | comparison | 7650 |

**Supplementary Table S2. Genetically nominated features (all four layers).** One row per nominated feature with stable identifier (UniProt / Ensembl gene / CpG / HMDB), the CKM outcome(s) it was nominated for, the nomination tier, and instrument details where available (n=1,875: proteome 84, transcriptome 81, methylome 1,395, metabolome 315). These counts are the deduplicated nominated features (with stable identifiers) across all eight CKM outcomes and nomination tiers — a different denominator from the per-representative-contrast counts in Table S1, so the two are not directly comparable feature-by-feature. The transcriptome set uses cis-MR p<0.05 and colocalization (the source is_causal field is unpopulated and was not used). Methylome instrument counts/F and transcriptome MR outcomes are not available at feature level on disk (NA). Provided as a machine-readable file.

*(Provided as a machine-readable supplementary file.)*

**Supplementary Table S3. Per-outcome nomination counts, metabolome (eGFR dominance).** Number of METABOLITES nominated for each CKM outcome — the metabolome is shown as representative, and the same dominance holds in every layer — illustrating that estimated glomerular filtration rate, the only quantitative trait and the best-powered GWAS, dominates the nominated set. Counts are per outcome, so a metabolite nominated for several outcomes is counted in each.

| **outcome** | **n_nominated** | **quantitative** |
| --- | --- | --- |
| T2D | 94 | no |
| NAFLD | 5 | no |
| CAD | 86 | no |
| AF | 36 | no |
| eGFR | 299 | yes |
| stroke | 8 | no |
| HF | 46 | no |
| CKD | 164 | no |

**Supplementary Table S4. Instrument diagnostics.** Median cis-instrument F-statistic, percentage with F>10, and percentage of multi-signal loci, for the proteome and metabolome (transcriptome and methylome feature-level cis-instrument F are not available; see Table S2). Metabolome multi-signal percentage is not defined — metabolites were clustered by shared metaboQTL gene — and is marked NA.

| **layer** | **medF** | **pctF10** | **pctMulti** |
| --- | --- | --- | --- |
| Proteome | 366.7 | 100 | 71.9 |
| Metabolome | 85.0 | 100 | NA |

**Supplementary Table S5a. Intervention estimand and ancestry per response cohort.**

| **Cohort** | **Intervention (class)** | **Estimand** | **Ancestry** |
| --- | --- | --- | --- |
| DiRECT | Caloric restriction (diet) | Within-person Δ (RCT active arm) | ~98% White (overall ITT) |
| By-Band-Sleeve | Bariatric surgery | Within-person Δ | White (British/Irish) 93%, Other 7% |
| STEP | Semaglutide (GLP-1RA) | Randomized treatment vs placebo | White 75.1%, Asian 12.4%, Black 5.0%, Other 7.6% |
| EMPEROR | Empagliflozin (SGLT2i) | Randomized treatment vs placebo (heart failure) | White ~85% |
| HERITAGE | Exercise training | Within-person Δ | European ~65% (n=424), African ~35% (n=230) |
| GSE83452 | Diet + bariatric | Within-person Δ | Not reported (single-center, Antwerp, Belgium) |
| GSE84046 | Energy restriction (high- vs normal-protein) | Within-person Δ (both randomized arms pooled) | Not reported (single-center, Wageningen, Netherlands) |
| DIRECT-PLUS | Mediterranean diet | Within-person Δ (RCT arm) | Not reported (single-center, Israel); 228 M / 28 F |
| PREDIMED (Arpón subset) | Mediterranean diet | Within-person Δ | Spanish (PREDIMED-Navarra, high-CV-risk) |
| Leipzig | Bariatric surgery | Within-person Δ | Not reported (single-center, Leipzig, Germany) |
| DRIFT2 | Behavioral | Within-person Δ | Not reported (source describes the cohort only as primarily non-Hispanic White) |
| Benton | Roux-en-Y gastric bypass | Within-person Δ | Not reported (single-center, Wellington, New Zealand) |
| Bagheri / Tanriverdi | Sleeve + RYGB (bariatric) | Within-person Δ | White 82.7%, African American 9.6%, Hispanic 7.7% |
| By-Band-Sleeve (Smith) | Bariatric surgery | Within-person Δ | White 86% |
| DiRECT (Corbin) | Caloric restriction (diet) | Within-person Δ | ~98% White (overall ITT) |

**Supplementary Table S5b. Source-overlap and ancestry audit.** QTL and outcome-GWAS sources, ancestry, and overlap with the response cohorts (overlap is minimal and, where present, biases two-sample MR toward observational association, making a null conservative).

| **component** | **sources** | **ancestry** | **overlap_note** |
| --- | --- | --- | --- |
| pQTL | UKB-PPP; deCODE | EUR | none with response cohorts; minor possible UKB↔outcome-GWAS overlap (inflates, not nulls) |
| eQTL | GTEx (via eQTL Catalogue: QTD000266, QTD000116) | EUR | none |
| mQTL | GoDMC | EUR | none |
| metaboQTL | Chen-CLSA | EUR | none |
| Outcome GWAS | 8 CKM GWAS (GCST accessions) | mostly EUR | minor possible UKB↔pQTL |
| Response cohorts | DiRECT,BBS,STEP,EMPEROR,DIRECT-PLUS,PREDIMED,DRIFT2,Benton,Bagheri,GSE83452/84046 | EUR/Western | — |

**Supplementary Table S6a. Methylome baseline-variance balance and matched effect size.** Per dataset and nomination tier: paired n, nominated and matched-comparison feature counts, raw permutation P, the variance-matched permutation P with its interquartile range, baseline-SD of nominated vs comparison CpGs and its Mann-Whitney P, OLS-adjusted P, and the matched Cliff's δ with 95% CI. The matched permutation P is the MEDIAN over 40 independent 5:1 matched draws, and the q25/q75 columns are the interquartile range across those draws: the statistic is sensitive to which comparison CpGs are drawn, so a single draw is not a property of the data.

| **dataset** | **tissue** | **intervention** | **n_pairs** | **tier** | **n_nominated** | **n_matched** | **raw_perm_p** | **matched_perm_p** | **matched_perm_p_q25** | **matched_perm_p_q75** | **baseSD_nominated** | **baseSD_comparison** | **baseSD_p** | **ols_p** | **cliffs_delta_matched** | **delta_lo** | **delta_hi** |
| --- | --- | --- | --- | --- | --- | --- | --- | --- | --- | --- | --- | --- | --- | --- | --- | --- | --- |
| DIRECT-PLUS | blood | Med-diet 18mo | 256 | mr_sig | 2608 | 13034 | 0.175 | 0.0455 | 0.0275 | 0.0747 | 0.0303 | 0.0321 | 2.3e-06 | 0.0103 | NA | NA | NA |
| DIRECT-PLUS | blood | Med-diet 18mo | 256 | smr_primary | 1531 | 7650 | 0.506 | 0.0652 | 0.0364 | 0.0983 | 0.0295 | 0.0321 | 2e-05 | 0.00427 | 0.031 | -0.003 | 0.064 |
| DIRECT-PLUS | blood | Med-diet 18mo | 256 | smr_heidi | 544 | 2720 | 0.175 | 0.803 | 0.652 | 0.917 | 0.0281 | 0.0321 | 4e-05 | 0.442 | NA | NA | NA |
| PREDIMED | blood | Med-diet 5yr | 36 | mr_sig | 2800 | 13995 | 0.743 | 0.278 | 0.158 | 0.317 | 0.0467 | 0.0493 | 1.8e-05 | 0.0955 | NA | NA | NA |
| PREDIMED | blood | Med-diet 5yr | 36 | smr_primary | 1639 | 8190 | 0.818 | 0.626 | 0.47 | 0.847 | 0.0475 | 0.0493 | 0.0043 | 0.161 | 0.007 | -0.026 | 0.039 |
| PREDIMED | blood | Med-diet 5yr | 36 | smr_heidi | 582 | 2905 | 0.515 | 0.785 | 0.711 | 0.871 | 0.0459 | 0.0493 | 0.015 | 0.145 | NA | NA | NA |
| Leipzig | blood | bariatric | 18 | mr_sig | 2607 | 13033 | 0.0193 | 0.241 | 0.163 | 0.354 | 0.0279 | 0.0303 | 1.7e-06 | 0.0229 | NA | NA | NA |
| Leipzig | blood | bariatric | 18 | smr_primary | 1530 | 7650 | 0.186 | 0.627 | 0.514 | 0.718 | 0.0278 | 0.0303 | 0.00011 | 0.233 | -0.009 | -0.044 | 0.025 |
| Leipzig | blood | bariatric | 18 | smr_heidi | 544 | 2720 | 0.877 | 0.674 | 0.527 | 0.872 | 0.0263 | 0.0302 | 0.00016 | 0.702 | NA | NA | NA |
| Benton | adipose | RYGB | 15 | mr_sig | 2599 | 12995 | 0.0196 | 0.637 | 0.443 | 0.739 | 0.0293 | 0.031 | 0.00021 | 0.853 | NA | NA | NA |
| Benton | adipose | RYGB | 15 | smr_primary | 1518 | 7590 | 0.0207 | 0.565 | 0.515 | 0.674 | 0.0284 | 0.031 | 0.0001 | 0.686 | -0.008 | -0.04 | 0.025 |
| Benton | adipose | RYGB | 15 | smr_heidi | 541 | 2705 | 0.282 | 0.765 | 0.64 | 0.875 | 0.0291 | 0.031 | 0.058 | 0.51 | NA | NA | NA |
| DRIFT2 | blood | behavioral 3mo | 62 | mr_sig | 2573 | 12863 | 0.274 | 0.81 | 0.636 | 0.888 | 0.0235 | 0.0248 | 0.00013 | 0.861 | NA | NA | NA |
| DRIFT2 | blood | behavioral 3mo | 62 | smr_primary | 1503 | 7513 | 0.482 | 0.805 | 0.688 | 0.919 | 0.0234 | 0.0247 | 0.0015 | 0.675 | -0.005 | -0.04 | 0.028 |
| DRIFT2 | blood | behavioral 3mo | 62 | smr_heidi | 536 | 2680 | 0.573 | 0.579 | 0.465 | 0.742 | 0.0214 | 0.0247 | 0.00024 | 0.711 | NA | NA | NA |

**Supplementary Table S6b. Proteome instrument-strength balance.** Per-instrument cis-F for nominated (n=90) vs comparison (n=1,595) SomaScan aptamer measurements, with summary balance statistics (standardized mean difference of raw and log F; Kolmogorov-Smirnov D and P) and UniProt/gene identifiers. Provided as a machine-readable file.

*(Provided as a machine-readable supplementary file.)*

**Supplementary Table S7. Restorative-direction (directional) results.** Per stratum: nominated restorative count/total and percentage, comparison percentage, odds ratio with 95% CI, exact binomial P vs 0.5, within-family Benjamini-Hochberg q (computed across these 15 directional strata; distinct from the manuscript multiplicity-table family-B definition), and Fisher P. Includes the EMPEROR (SGLT2i) and HERITAGE (exercise) strata. For two-group strata, the odds-ratio Wald P reported in the main text (e.g. HERITAGE OR 1.64, P = 0.18) corresponds to the OR and its 95% CI; the binomial-vs-0.5 and Fisher P columns are complementary one-group and exact tests. The margin-preserving permutation (Methods) is the primary directional test; the binomial-vs-0.5 and odds-ratio columns are descriptive secondary summaries.

| **stratum** | **k_nominated** | **n_nominated** | **nominated_pct** | **k_comparison** | **n_comparison** | **comparison_pct** | **OR** | **OR_lo** | **OR_hi** | **binom_vs50_P** | **fisher_P** | **binom_q_BH** | **test_type** |
| --- | --- | --- | --- | --- | --- | --- | --- | --- | --- | --- | --- | --- | --- |
| Proteome Olink coloc | 14 | 20 | 70 | NA | NA | 49 | 2.39 | 0.91 | 6.27 | 0.115 | 0.076 | 0.456 | one-group vs 0.5 |
| Proteome Olink MR | 38 | 74 | 51 | 493 | 993 | 50 | 1.07 | 0.67 | 1.72 | 0.908 | 0.777 | 1.0 | two-group OR |
| Proteome SomaScan MR | 48 | 84 | 57 | 525 | 1121 | 47 | 1.51 | 0.97 | 2.37 | 0.23 | 0.071 | 0.456 | two-group OR |
| Transcriptome liver MR | 24 | 56 | 43 | 613 | 1266 | 48 | 0.8 | 0.47 | 1.37 | 0.35 | 0.416 | 0.583 | two-group OR |
| Transcriptome liver coloc | 12 | 25 | 48 | NA | NA | 48 | 0.99 | 0.45 | 2.19 | 1.0 | 0.985 | 1.0 | one-group vs 0.5 |
| Metabolome Bagheri (bariatric) | 86 | 164 | 52 | NA | NA | 44 | 1.41 | 0.91 | 2.18 | 0.585 | 0.151 | 0.739 | one-group vs 0.5 |
| Metabolome BBS (bariatric) | 128 | 255 | 50 | NA | NA | 46 | 1.19 | 0.85 | 1.67 | 1.0 | 0.306 | 1.0 | one-group vs 0.5 |
| Metabolome DiRECT (diet) | 93 | 207 | 45 | NA | NA | 46 | 0.96 | 0.66 | 1.41 | 0.164 | 0.923 | 0.456 | one-group vs 0.5 |
| Methylome DIRECT-PLUS | 796 | 1531 | 52 | NA | NA | 50 | 1.08 | 0.97 | 1.19 | 0.125 | 0.146 | 0.456 | one-group vs 0.5 |
| Methylome PREDIMED | 779 | 1639 | 48 | NA | NA | 50 | 0.9 | 0.82 | 1.0 | 0.048 | 0.044 | 0.405 | one-group vs 0.5 |
| Methylome Leipzig | 776 | 1530 | 51 | NA | NA | 50 | 1.03 | 0.93 | 1.14 | 0.591 | 0.544 | 0.739 | one-group vs 0.5 |
| Methylome Benton(adipose) | 721 | 1518 | 47 | NA | NA | 50 | 0.9 | 0.82 | 1.0 | 0.054 | 0.052 | 0.405 | one-group vs 0.5 |
| Methylome DRIFT2 | 763 | 1503 | 51 | NA | NA | 50 | 1.03 | 0.93 | 1.14 | 0.57 | 0.561 | 0.739 | one-group vs 0.5 |
| Proteome EMPEROR (SGLT2i, MR) | 40 | 69 | 58 | NA | NA | 50 | NA | NA | NA | 0.228 | 0.262 | 0.456 | one-group vs 0.5 |
| Proteome HERITAGE (exercise) | 22 | 36 | 61 | 92 | 188 | 49 | 1.64 | 0.79 | 3.4 | 0.243 | 0.205 | 0.456 | two-group OR (OR-Wald P=0.18) |

**Supplementary Table S8a. Cross-platform nominated proteins.** Proteins nominated on both Olink/UKB-PPP and SomaScan/deCODE, and whether each responded preferentially.

| **cross_platform_protein** | **nominated_by** | **preferentially_responsive** |
| --- | --- | --- |
| TNFSF12 | Olink+SomaScan | no |
| APOH | Olink+SomaScan | no |
| ARG1 | Olink+SomaScan | no |
| MMP12 | Olink+SomaScan | no |
| LRIG1 | Olink+SomaScan | no |
| IDI2 | Olink+SomaScan | no |
| SPON1 | Olink+SomaScan | no |

**Supplementary Table S8b. Cross-layer node-bridge matrix.** Full member-gene lists (no ellipsis) for every pairwise, three-way and four-way overlap among the 74 multi-layer reversal-tested genetically-nominated nodes (HGNC symbols). The node universe is the set of genes nominated for any non-eGFR CKM outcome and reversal-tested in at least two layers; 'NA' denotes no shared genes. Provided as a single-header, tab-separated, machine-readable file.

| **layer_pair** | **n_overlap** | **genes** |
| --- | --- | --- |
| proteome∩transcriptome | 5 | LRRC37A2,NFATC1,NMB,PDCD5,SORT1 |
| methylation∩transcriptome | 17 | CELSR2,CWF19L1,CWH43,DNA2,LAMA5,MAN2C1,NFATC1,PGAP3,PSRC1,SHROOM3,SLC22A3,SLC2A4RG,SORT1,SYPL2,TFDP2,TMEM60,ZNF436 |
| methylation∩proteome | 35 | ACP1,APOA5,B3GNT8,CALCOCO2,CDKN1A,CTSS,DPEP1,ECM1,FGFR4,FN1,FOXO3,FURIN,GATM,GCKR,IL6R,INHBC,ITIH1,ITIH4,ITPKA,MMP12,NAGLU,NFATC1,NOTCH2,PCSK9,PDE5A,QSOX2,RNF43,SEMA5A,SNUPN,SORT1,STAT3,TCEA2,TGFB1,TIMP2,VAMP8 |
| metabolite∩methylation | 15 | ALMS1,ATXN2,CDK12,DNAJC16,DPEP1,GATM,GCKR,GSTA2,IGF2R,KDM5A,NAT2,PEMT,SLC22A2,SLC47A1,SLC7A9 |
| metabolite∩proteome | 10 | AOC1,ARG1,BLMH,COMT,CRAT,DPEP1,GATM,GCKR,IVD,NT5C3A |
| metabolite∩transcriptome | 2 | PPDPF,SLC17A1 |
| 3way:proteome∩transcriptome∩methylation | 2 | NFATC1,SORT1 |
| 3way:metabolite∩proteome∩methylation | 3 | DPEP1,GATM,GCKR |
| 3way:metabolite∩transcriptome∩methylation | 0 | NA |
| 3way:metabolite∩proteome∩transcriptome | 0 | NA |
| 4way:all | 0 | NA |

**Supplementary Table S9. Software, package versions, random seeds, and code/data availability.**

| **component** | **version** |
| --- | --- |
| python | 3.12.4 |
| pandas | 2.3.3 |
| numpy | 2.0.1 |
| scipy | 1.14.0 |
| R | 4.4.1 |
| R: minfi | 1.52.1 |
| R: EpiDISH | 2.22.0 |
| R: coloc | 5.2.3 |
| R: TwoSampleMR | 0.7.8 |
| R: ggplot2 | 3.5.1 |
| R: cowplot | 1.1.3 |
| R: limma | 3.62.2 |
| R: data.table | 1.15.4 |
| random seed | 42 (numpy default_rng; methylation/metabolome contrast + figure engines); metabolite_mr_robust.py = seed 7 |
| code repository | https://github.com/badboybert/ckm-4layer-null |
| archive DOI | 10.5281/zenodo.20965271 |
| OS | Windows-11-10.0.26200-SP0 |

**Supplementary Table S10a. Cell-composition sensitivity of the methylome contrast.** Variance-matched permutation P for nominated vs comparison CpGs, before (matchP_unadj) and after (matchP_celladj) adjusting each CpG's within-person change for the within-person change in EpiDISH-estimated blood cell-type proportions, for the three blood cohorts under both the eGFR-inclusive and leave-eGFR-out nominated sets, with within-config Benjamini-Hochberg q. No tier survives FDR in either configuration. (For DIRECT-PLUS and Leipzig the recomputed unadjusted per-CpG changes agree with the deposited reversal tables to within 3 × 10⁻⁸, the deposited files' stored precision; for DRIFT2 about a fifth of probes carry no paired value owing to QC/pairing, and its contrasts are likewise non-significant.)

| **cohort** | **config** | **tier** | **matchP_unadj** | **matchP_celladj** | **q_celladj** |
| --- | --- | --- | --- | --- | --- |
| directplus | eGFR_inclusive | mr_sig | 0.0525 | 0.212 | 0.636 |
| directplus | eGFR_inclusive | smr_sig | 0.128 | 0.0532 | 0.5352 |
| directplus | eGFR_inclusive | smr_primary | 0.0682 | 0.169 | 0.636 |
| directplus | eGFR_inclusive | smr_heidi | 0.865 | 0.0892 | 0.5352 |
| directplus | leave_eGFR_out | mr_sig | 0.001 | 0.303 | 0.7272000000000001 |
| directplus | leave_eGFR_out | smr_sig | 0.0132 | 0.154 | 0.616 |
| directplus | leave_eGFR_out | smr_primary | 0.0137 | 0.454 | 0.8115000000000001 |
| directplus | leave_eGFR_out | smr_heidi | 0.324 | 0.208 | 0.624 |
| leipzig | eGFR_inclusive | mr_sig | 0.275 | 0.817 | 0.8169999999999998 |
| leipzig | eGFR_inclusive | smr_sig | 0.713 | 0.743 | 0.8169999999999998 |
| leipzig | eGFR_inclusive | smr_primary | 0.588 | 0.34 | 0.8160000000000001 |
| leipzig | eGFR_inclusive | smr_heidi | 0.688 | 0.52 | 0.8169999999999998 |
| leipzig | leave_eGFR_out | mr_sig | 0.424 | 0.045 | 0.54 |
| leipzig | leave_eGFR_out | smr_sig | 0.592 | 0.135 | 0.616 |
| leipzig | leave_eGFR_out | smr_primary | 0.689 | 0.53 | 0.8115000000000001 |
| leipzig | leave_eGFR_out | smr_heidi | 0.117 | 0.541 | 0.8115000000000001 |
| drift2 | eGFR_inclusive | mr_sig | 0.809 | 0.669 | 0.8169999999999998 |
| drift2 | eGFR_inclusive | smr_sig | 0.462 | 0.755 | 0.8169999999999998 |
| drift2 | eGFR_inclusive | smr_primary | 0.549 | 0.807 | 0.8169999999999998 |
| drift2 | eGFR_inclusive | smr_heidi | 0.665 | 0.669 | 0.8169999999999998 |
| drift2 | leave_eGFR_out | mr_sig | 0.263 | 0.818 | 0.818 |
| drift2 | leave_eGFR_out | smr_sig | 0.22 | 0.63 | 0.818 |
| drift2 | leave_eGFR_out | smr_primary | 0.261 | 0.791 | 0.818 |
| drift2 | leave_eGFR_out | smr_heidi | 0.802 | 0.686 | 0.818 |

**Supplementary Table S10b. Mean within-person change in estimated blood cell-type fractions** (post - pre) per cohort, from EpiDISH reference-based deconvolution.

| **cohort** | **cell_type** | **mean_delta** | **sd_delta** | **n_pairs** |
| --- | --- | --- | --- | --- |
| directplus | B | 0.00108 | 0.01013 | 256 |
| directplus | NK | 0.00818 | 0.02532 | 256 |
| directplus | CD4T | 0.00577 | 0.03368 | 256 |
| directplus | CD8T | 0.00612 | 0.03148 | 256 |
| directplus | Mono | 0.00074 | 0.01504 | 256 |
| directplus | Neutro | -0.02217 | 0.06572 | 256 |
| directplus | Eosino | 0.00028 | 0.0021 | 256 |
| leipzig | B | -0.00352 | 0.02018 | 18 |
| leipzig | NK | 0.00389 | 0.03546 | 18 |
| leipzig | CD4T | 0.02244 | 0.06442 | 18 |
| leipzig | CD8T | -0.00574 | 0.02446 | 18 |
| leipzig | Mono | -0.00269 | 0.03973 | 18 |
| leipzig | Neutro | -0.01439 | 0.15039 | 18 |
| leipzig | Eosino | 0.0 | 0.0 | 18 |
| drift2 | B | 0.00225 | 0.01308 | 62 |
| drift2 | NK | 0.00027 | 0.02015 | 62 |
| drift2 | CD4T | 0.00319 | 0.04505 | 62 |
| drift2 | CD8T | 0.00867 | 0.0334 | 62 |
| drift2 | Mono | 0.0043 | 0.01987 | 62 |
| drift2 | Neutro | -0.01871 | 0.08562 | 62 |
| drift2 | Eosino | 2e-05 | 0.00081 | 62 |

**Supplementary Table S11. Master analysis specification.** Every analysis with its layer, dataset/cohort, intervention class, tissue, participant N, nominated/comparison feature counts, estimand, response metric, nomination tier, comparator rule, dependence/resampling unit, effect estimate (95% CI; permutation P) and pre-specified or post-hoc status. Machine-readable in Supplementary Data 1.

**Supplementary Table S12. Multiplicity table.** Every statistical test with its family, raw P, Benjamini-Hochberg q within that family, pre-specified or post-hoc status, and whether it survives false-discovery control at q < 0.05. Machine-readable in Supplementary Data 1.

### **Supplementary Figure legends**

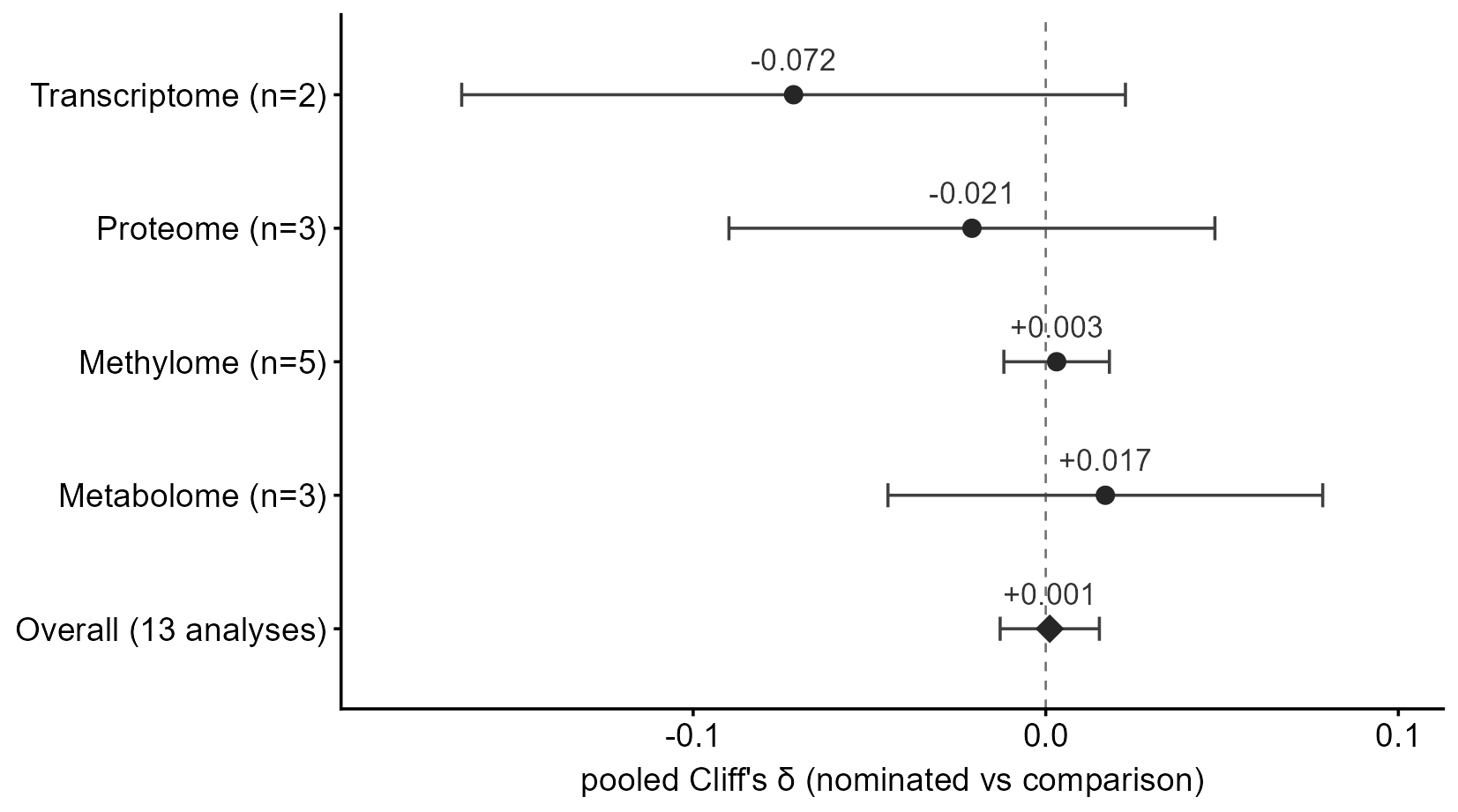

**Supplementary Figure S1. Random-effects subgroup synthesis.** Pooled Cliff's δ overall and by molecular layer, all within the small-effect reference region (±0.10).

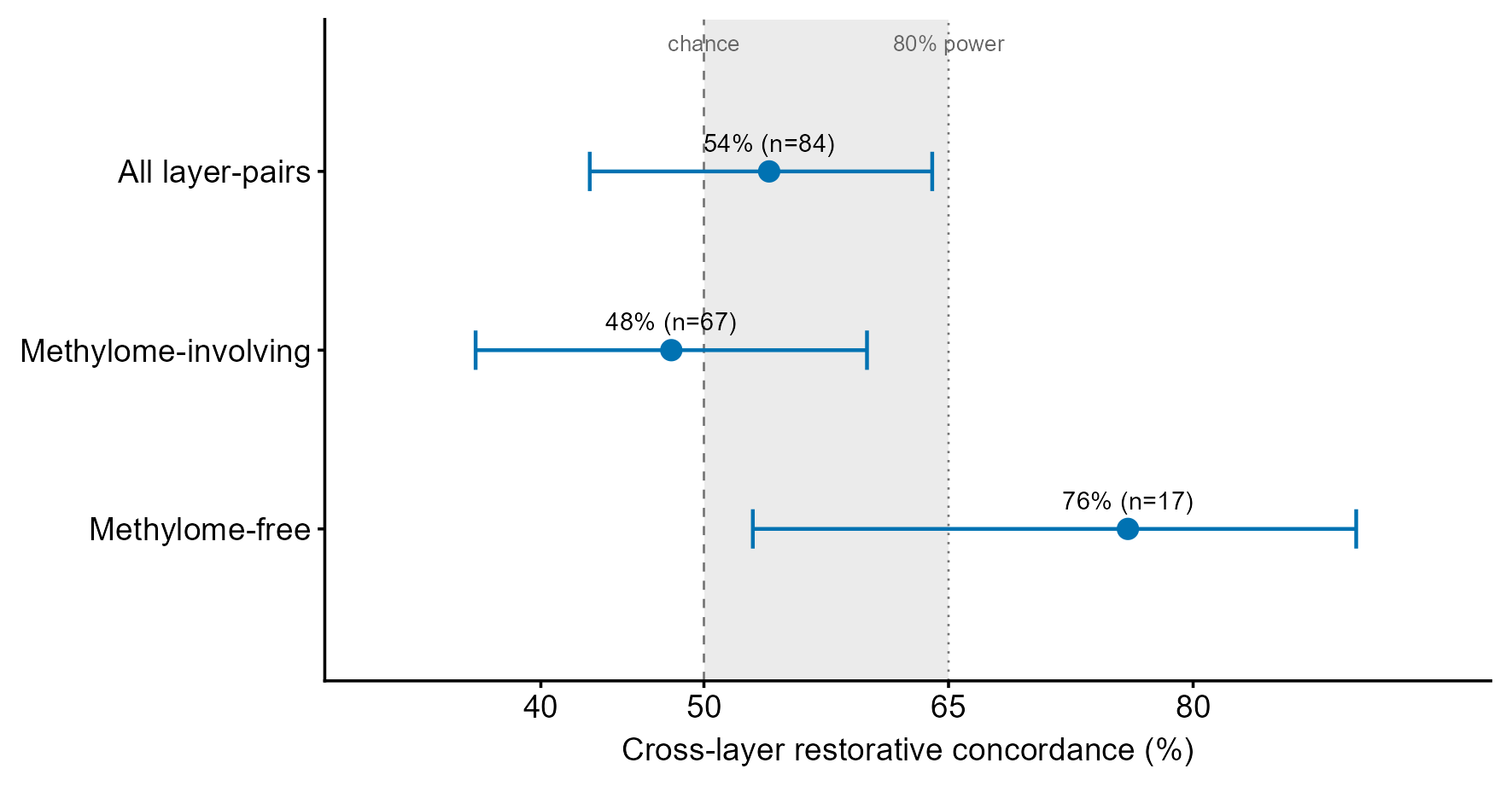

**Supplementary Figure S2. Cross-layer node concordance.** 45/84 (54%) of layer-pairs concordant in restorative direction, indistinguishable from the within-node permutation expectation and underpowered; hypothesis-generating only.

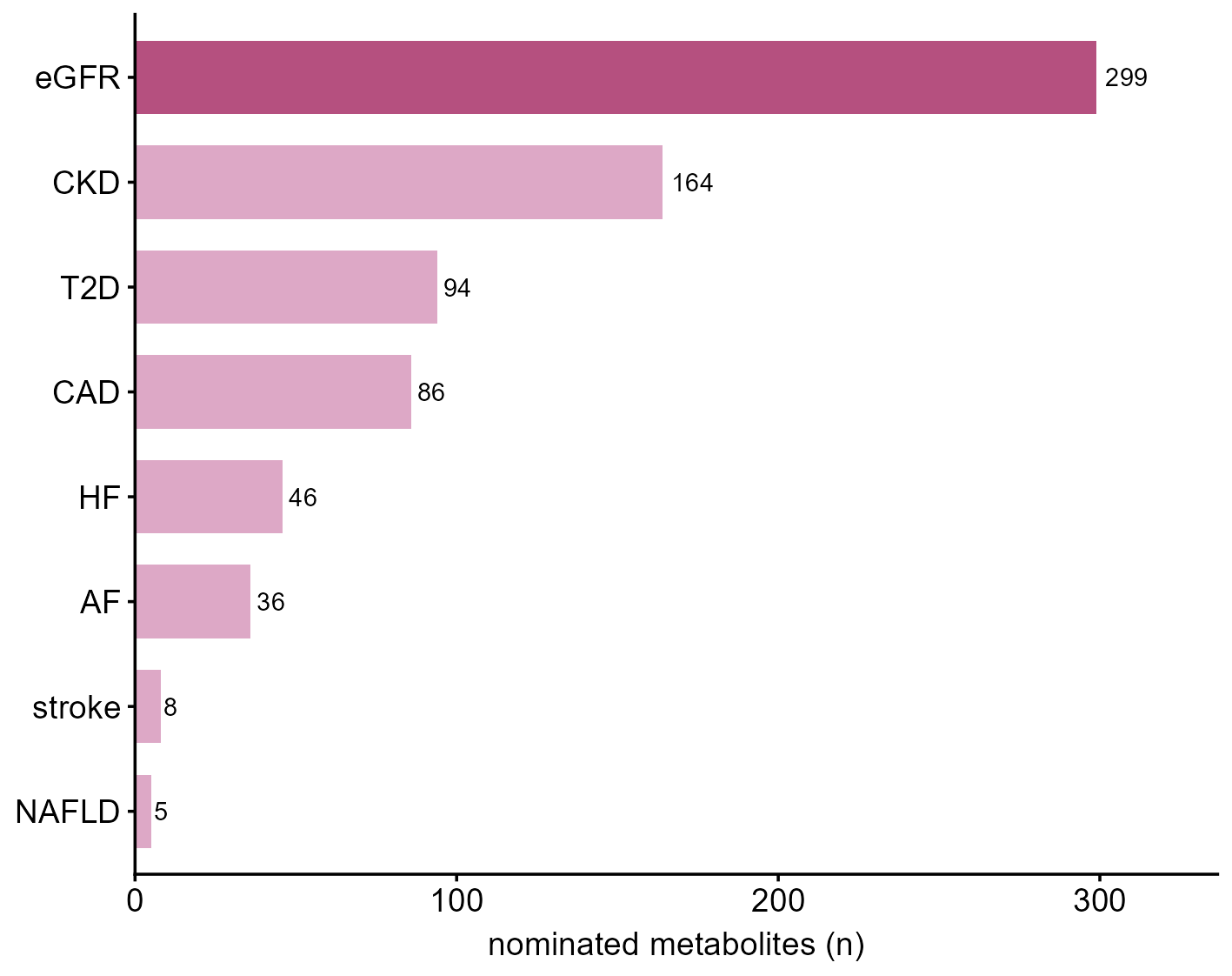

**Supplementary Figure S3. eGFR-set dominance in the metabolome.** Per-outcome nominated-set size for the METABOLOME, shown as representative of every layer (the panel's axis is labeled accordingly), showing that eGFR — the only quantitative outcome — dominates the nominated set and motivating the pre-specified leave-eGFR-out primary analysis.

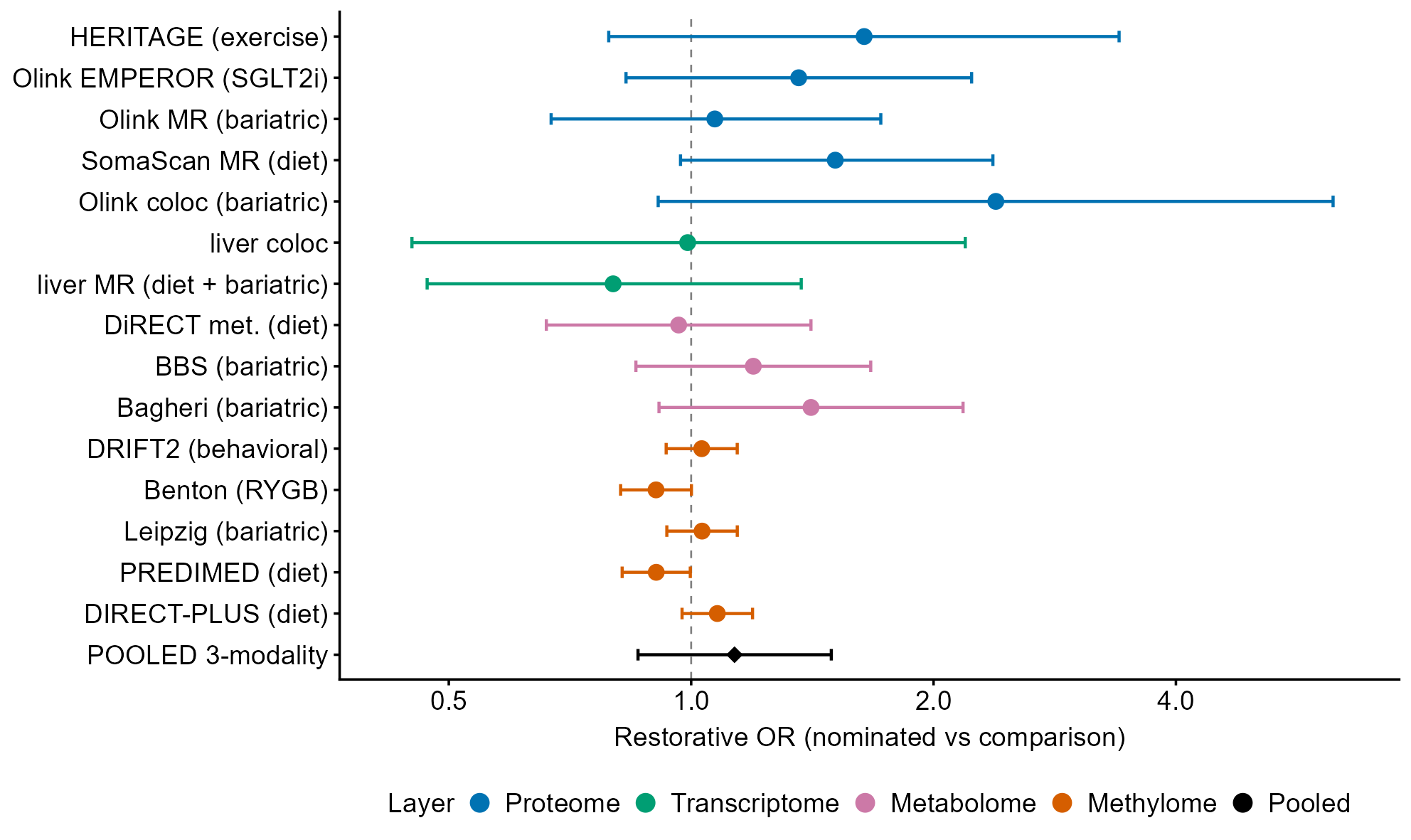

**Supplementary Figure S4. Exploratory directional analysis.** Restorative-direction forest (per-layer and pooled odds ratios; one interval marginally excludes 1, PREDIMED at odds ratio 0.90, 95% CI 0.82 to 0.998, and no stratum is significant after false-discovery control, minimum q = 0.41); the margin-preserving permutation results are reported in the Results and Methods; inconclusive.

### **Figure alt text (accessibility)**

- **Figure 1.** Four-stage study-design schematic (molecular layers -> genetic nomination -> nominated vs comparison groups -> molecular response), a log-scale feature-flow chart per layer, and a calibration panel showing canonical causal proteins recovered.
- **Figure 2.** Four panels: (a) forest of Cliff's δ (nominated vs comparison response magnitude) with 95% CIs for all 13 analyses, colored by layer, against a ±0.10 reference band, pooled estimate near zero; (b) pooled δ per intervention class (single-analysis classes shown as points, not pooled); (c) magnitude-by-direction scatter clustering at the null origin; (d) label-permutation null distributions for representative proteome/transcriptome/metabolome cells with the observed delta within the null.
- **Figure 3.** Violin-and-box plots of per-feature response magnitude for nominated vs comparison features, one representative dataset per layer; distributions overlap.
- **Figure 4.** Five-panel methylome confound analysis: (a) baseline-SD density (nominated lower-variance) in two cohorts; (b) permutation P rising from raw to adjusted; (c) island/promoter enrichment odds ratios; (d) variance-matched permutation P across cohorts x tiers, 14 of 15 above 0.05 (DIRECT-PLUS at the MR-significant tier is the exception); (e) leave-eGFR-out permutation P before vs after blood cell-composition adjustment, showing the single-cohort (DIRECT-PLUS) signal rising above 0.05 after adjustment.
- **Figure 5.** Robustness panels: permutation P across nomination-stringency tiers; cis-instrument F distributions; baseline instrument-strength balance; metabolite gene-clustering effect on intervals.
- **Supplementary Figures S1-S4.** As described in the legends above.
